# Inflammation and nitro-oxidative stress in current suicidal attempts and current suicidal ideation: a systematic review and meta-analysis

**DOI:** 10.1101/2021.09.09.21263363

**Authors:** Asara Vasupanrajit, Ketsupar Jirakran, Chavit Tunvirachaisakul, Marco Solmi, Michael Maes

## Abstract

A meta-analysis showed a significant association between activated immune-inflammatory and nitro-oxidative (IO&NS) pathways and suicide attempts (SA). There is no data on whether suicidal ideation (SI) is accompanied by activated IO&NS pathways and whether there are differences between SA and SI. The current study searched PubMed, Google Scholar, and Web of Science, for articles published from inception until May 10, 2021, and systematically reviewed and meta-analyzed the association between recent SA/SI (< 3 months) and IO&NS biomarkers. We included studies which compared psychiatric patients with and without SA and SI and controls (either healthy controls or patients without SA or SI) and used meta-analysis (random-effect model with restricted maximum-likelihood) to delineate effect sizes with 95% confidence intervals (CI). Our search included 59 studies comprising 4.034 SA/SI cases and 12.377 controls. Patients with SA/SI showed activated IO&NS pathways (SMD: 0.299; CI: 0.200; 0.397) when compared to controls. The immune profiles were more strongly associated with SA than with SI, particularly when compared to healthy controls, as evidenced by activated IO&NS (SMD: 0.796; CI: 0.503; 1.089), immune (SMD: 1.409; CI: 0.637; 1.462), inflammatory (SMD: 1.200; CI: 0.584; 1.816), and neurotoxic (SMD: 0.904; CI: 0.431; 1.378) pathways. The effects sizes of the IO&NS, immune and inflammatory profiles were significantly greater in SA than in SI. In conclusion: increased neurotoxicity due to inflammation and nitro-oxidative stress and lowered neuroprotection explains at least in part why psychiatric patients show increased SA and SI. The IO&NS pathways are more pronounced in recent SA than in SI.

## Introduction

Non-fatal suicide behaviors (SB) are self-injurious behaviors, which may be classified according to their severity into suicidal ideation (SI) and suicide attempts (SA)^1^. A study across 17 countries shows that SI has a greater lifetime prevalence than SA, namely 9.2% and 2.7%, respectively^2^, indicating that not all individuals with SI will attempt suicide. Many different risk factors may lead to SB^3, 4^ but psychiatric disorders are the major risk factors^5, 6^. Individuals with major depressive disorder (MDD) show an almost eight-fold increased risk of suicide, whilst bipolar disorder (BD) and schizophrenia (SCZ) show a six-fold increased risk as compared with individuals without these psychiatric disorders^6^.

There is now evidence that activation of immune-inflammatory and nitro-oxidative (IO&NS) pathways is involved in the pathophysiology of major psychiatric disorders, including MDD, BD, and SCZ^7-10^. These disorders are characterized by a simultaneous activation of the immune-inflammatory response system (IRS) and the compensatory immune-regulatory system (CIRS), which may down-regulate the IRS. **Electronic Supplementary File (ESF) 1 Table 1** shows the IRS-CIRS pathways, and their features involved in those major psychiatric disoders.^11-13^. IRS activation is accompanied by the induction of nitro-oxidative (O&NS) pathways with increased production of reactive oxygen (ROS) and nitrogen species (RNS) especially when the antioxidant defenses are reduced^7^. The consequent redox disbalance may cause increased damage to lipids, proteins, DNA, and mitochondria^7^. Moreover, many IO&NS markers have neurotoxic effects, including interleukin (IL-)2, IL-6, tumor necrosis factor-alpha (TNF-α), interferon-gamma (IFN-γ), some chemokines (i.e., CCL-2, CCL-5), C-reactive protein (CRP), quinolinic acid (QA), picolinic acid (PA), malondialdehyde (MDA), and nitric oxide metabolites (NOx)^13, 14^. Increased neurotoxicity may cause dysfunctions in gray and white matter plasticity, which in turn are associated with affective disorders and SCZ^13-15^. In contrast, antioxidants, including albumin, vitamin-D, and high-density lipoprotein (HDL), and neurotrophic substances, including brain-derived neurotrophic factor (BDNF)^16^, may have neuroprotective effects thereby protecting the central nervous system (CNS) against the neurotoxic effects of the IRS^7^.

Since the 1990ts, there were also studies implicating IO&NS pathways in the pathophysiology of SB^17, 18^. Since then, a rising number of studies have examined the relationship between SB (either SA or SI) and IO&NS biomarkers, including IL-1β, IL-6, TNF-α, IFN-γ, chemokines such as IL-8, and the neutrophil-to-lymphocyte ratio; CIRS markers including IL-4, IL-10, and soluble IL-2 receptor (sIL-2R); acute phase response (APR) productions such as CRP and albumin; O&NS markers (i.e., MDA, NOx); and antioxidants or neurotrophic factors including albumin, BDNF, omega-3 polyunsaturated fatty acids (PUFAs), HDL-cholesterol, total antioxidant capacity (TAC), and vitamin D3^19,23,24^. The increase in IL-6 in cerebrospinal fluid (CSF), blood, and postmortem brain is one of the most consistently reported abnormalities in SB, while other pro-inflammatory cytokines showed more controversial results ^20, 21^. It is noteworthy that the cytokine imbalance in SB may present with activation of the tryptophan catabolite (TRYCAT) pathway leading to lowered levels of tryptophan (TRP) and increased levels of TRYCATs including QA, PA, and kynurenine (KYN)^4, 22, 23^. Meta-analyses, which examined associations with cytokines other than IL-6, frequently reported controversial findings. For example, Black and Miller^24^ discovered significantly higher IL-1β blood levels, whereas Ducasse et al.^25^ found a medium effect size with lower IL-2 and IL-4 plasma but higher TGF-β plasma levels. CRP (positively)^26, 27^ and BDNF (negatively)^28^ were significantly associated with SB, while SB was associated with BDNF in plasma^28^ but not in serum^28, 29^.

Importantly, most previous meta-analyses on IO&NS blood biomarkers and suicide examined SA and SI patients lumped together into one SB group without considering possible differences between these groups^24-26^. Hence, the current study aimed to systematically review and meta-analyze the association between IO&NS blood biomarkers and recent SB and to delineate differences among SA and SI. The examination of solitary IO&NS biomarkers is less relevant because these compounds take part in highly connected networks and subnetworks. In this regard, Maes et al.^9, 15^ showed that the combination of several IO&NS biomarkers into weighted composite scores may improve our understanding of the pathways involved in affective disorders, schizophrenia and SB. Therefore, we here examine whether SA and/or SI are associated with composite scores reflecting IO&NS (the primary analyses) and its subdomains namely IRS, inflammation, neurotoxicity, and neuroprotection (the secondary analyses) as described in ESF1 Table 1. In addition, we also examine solitary biomarkers such as CRP (part of the inflammation profile) and BDNF (part of neuroprotection profile). Based on previous knowledge, we hypothesized that a) in patients with SA and SI, IO&NS, and IRS, inflammation, neurotoxicity, and CRP are increased, while neuroprotection and BDNF are reduced when compared with controls; and b) that aberrations in these functional profiles are more pronounced in SA than in SI.

**Table 1.**
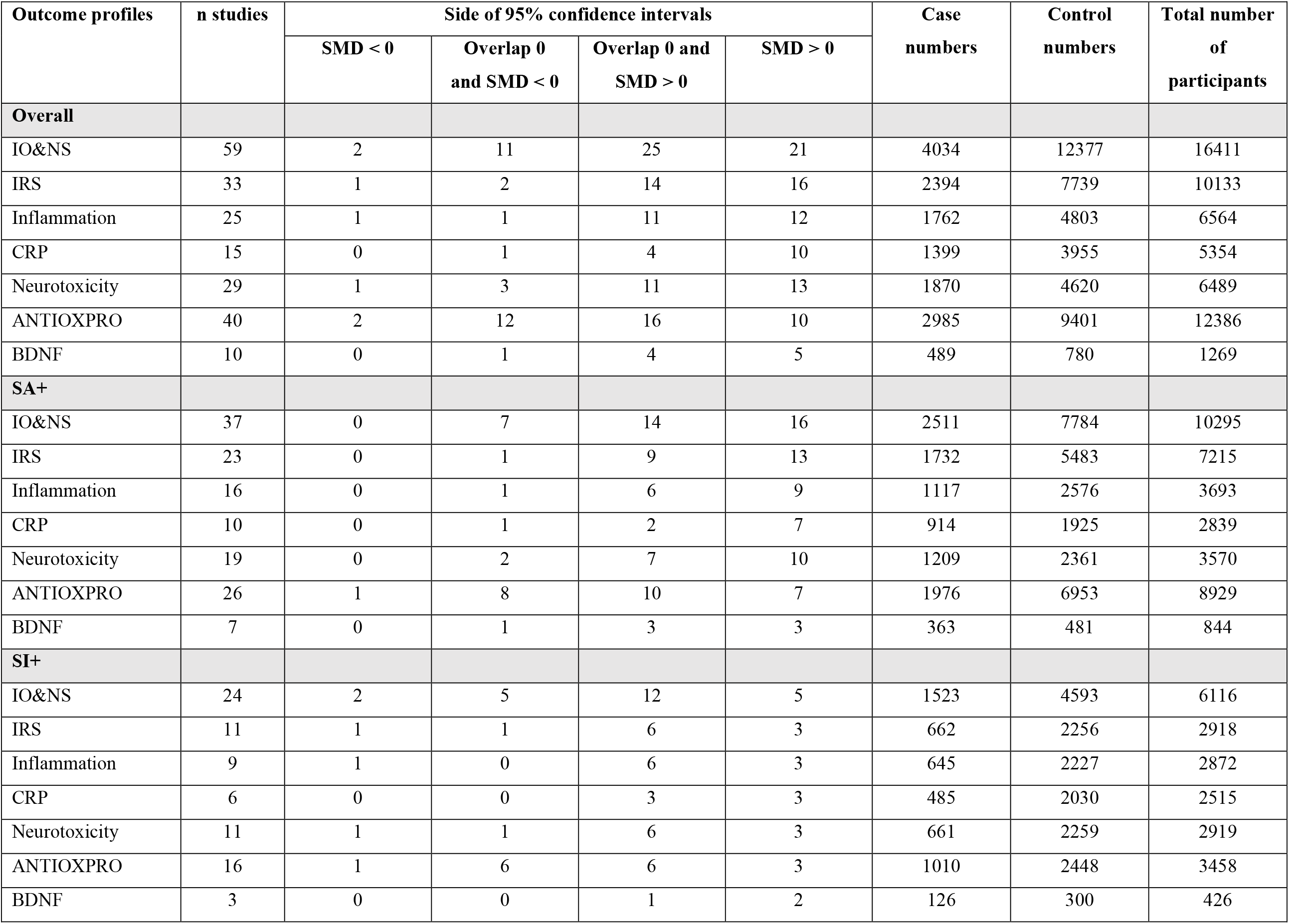
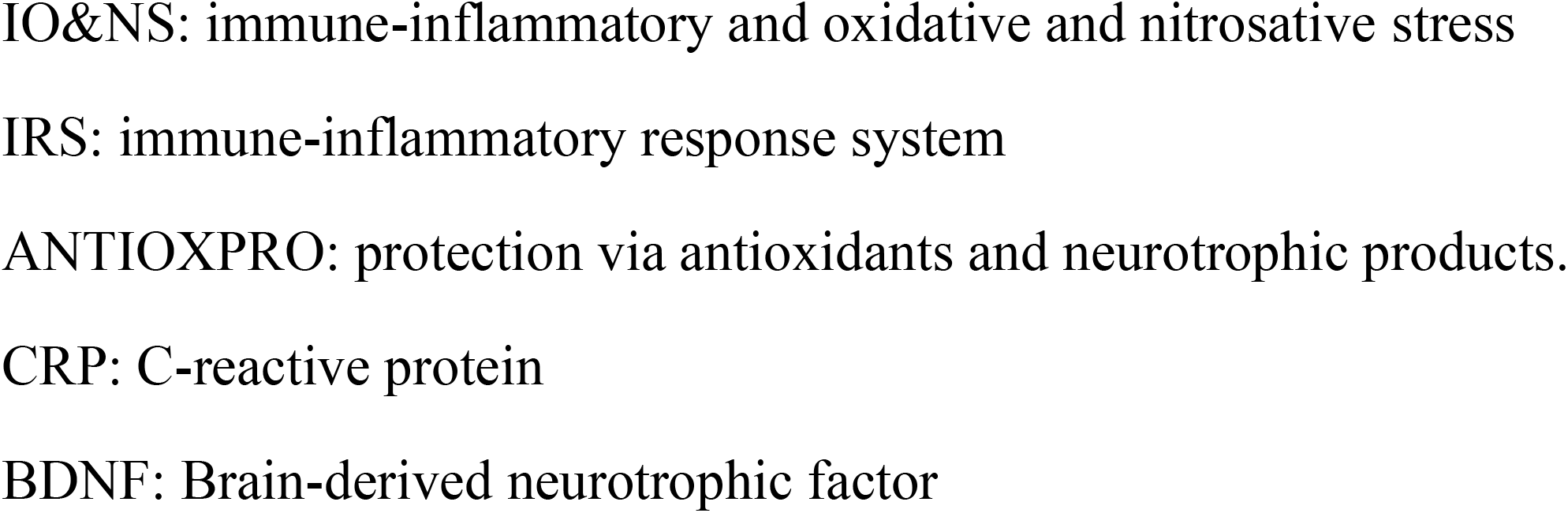
Number of cases with suicidal attempts (SA+), suicidal ideation (SI+) and controls in the different meta-analyses and side of standardized mean difference (SMD) and the 95% confidence intervals with respect to zero SMD.

## Materials and methods

Our methodological approach was based on the guidelines of the Preferred Reporting Items for Systematic Reviews and Meta-Analyses (PRISMA) 2020^30^, the Cochrane Handbook for Systematic Reviews and Interventions^31^, and the Meta-Analyses of Observational Studies in Epidemiology (MOOSE)^32^. We followed an a priori protocol which is available upon request from the last author. The draft or process of this study did not directly or indirectly involve public or patient representatives.

### Scope of systematic review

This study classified SB into 2 groups, namely SA and SI^1^. We combined different biomarkers into composites according to their well-established functions (ESF 1 Table 1). This study began by selecting articles published in peer-reviewed journals and then searched further records in the studies’ reference lists and grey literature. We included English case-control studies or cohort studies when these included a control group and compared SB (SA+ or SI+ within 3 months before the study) with controls, including non-SB patients (SA- or SI-) and/or healthy controls (HC), alone and together. As such, we also examined differences in effect sizes between SA+ *versus* SA- and SI+ *versus* SI-within patients with psychiatric disorders. Moreover, we also examined possible differences in effect sizes between SA and SI. We examined IO&NS biomarkers in blood, serum, and plasma but not CSF, urine, platelets, or stimulated whole blood in articles that included patients and controls of all sexes, all ages, and ethnicities.

We approached the authors of eligible studies which did not publish mean and standard deviation (SD) or standard error (SE) data and requested for means, SD, and the number of cases and controls. We calculated mean and SD values when the authors did not answer to our request, using formulae provided by Wan et al.^33^ for studies reporting median with either interquartile range (IQR) or minimum/maximum values. We excluded studies that reported on animal models, translational and genetic research, and studies that did not include controls. Any reasons for the exclusion of research were recorded.

### Literature search and data extraction

A systematic review of studies was performed utilizing electronic databases including PUBMED/MEDLINE, Google Scholar, and Web of Science from inception to 10th May 2021. The search used the major terms of IO&NS biomarkers and suicide **(ESF 1 Table 2)**, include “inflamm*” OR immun*”, “cytokine”, “chemokine”, “IL-6”, “IL-1”, “interleukin”, “C-Reactive Protein”, “CRP”, “tumor necrosis factor”, “TNF”, “interferon”, “IFN”, “Transforming growth factor”, “TGF”, “Tryptophan”, “Oxidative stress”, “Antioxidants”, “Zinc”, “Vitamin”, “Albumin”, “Nitric oxide”, “Lipid hydroperoxides”, “Omega 3”, “Coenzyme Q10”, “DHA”, “suicid*”. We manually searched the reference lists of the included studies along with prior meta-analysis studies.

The first author (AV) screened the eligibility manuscripts depending on the titles and abstracts, then obtained the full text of the potentially eligible studies and extracted the relevant data into a predefined Excel spread sheet. The second author (KJ) independently checked the extracted data once the first author (AV) completed all information. In case of disagreement, the last author (MM) was consulted. We used a methodological quality and red-point score checklist, namely the immune cofounder’s scale (ICS), adapted from Andrés-Rodríguez et al.^34^ and Vasupanrajit et al.^19^, which was slightly modified by the last author **(ESF 1 Table 3)**.

### Statistical Analysis

This study performed statistical analysis using the Comprehensive meta-analysis (CMA) V3 software. The precise methods were described previously.^18^ The primary outcome was the pooled standardized mean difference (SMD) of the IO&NS profiles. The secondary outcomes were pooled SMD of the IO&NS subdomains, namely IRS, inflammation, CRP, neurotoxicity, antioxidant/neuroprotective (ANTIOXPRO), and BDNF, which were examined to determine whether components of the IO&NS are more significant to recent SA and SI. Hence, in the present MA, we employed synthetic scores which reflect outcome profiles and compared these scores between SA and SI and controls. For studies that reported more than one IO&NS biomarker, a synthetic score was calculated by computing the mean of the outcomes and assuming dependence. We used a random effect model with restricted maximum-likelihood because there are differences in study design, measuring time, and participants’ characteristics across studies. The random-effects subgroup meta-analysis were performed, comparing patient subgroups (either SA+ or SI+) *versus* controls. This study computed the SMD values with 95% confidence intervals (CI) and used an alpha level of 0.05 indicating statistically significant results (two sided tests). The Cochrane Q test and the *I*^*2*^ metric were computed, but Tau (τ) and τ^2^ was used to evaluate heterogeneity^35, 36^. If τ^2^ was imprecise, we evaluated possible sources of heterogeneity across studies when at least ten studies presented data of the same profile, using either subgroup meta-analysis (with a minimum of three studies per subgroup) or random-effects meta-regression analyses (with minimum of ten studies).

The leave-one-out method was used to conduct sensitivity analyses to examine the robustness of the pooled combined meta-analysis effects and between-study heterogeneity. To assess small study effects, including publication bias, we employed the classic fail-safe N method, Kendall tau with continuity correction (using one-tailed p-values), and Egger’s regression intercept (using one-tailed p-values). We also employing funnel plots, which display study precision on the y-axis and the SDM on the x-axis, to detect small study effects or systemic heterogeneity by simultaneously presenting the observed studies and the imputed missing values.

## Results

### Search results

Our search strategy identified 2451 reports. Of those, 64 articles^17, 18, 20, 37-97^ were assessed in our systematic review after removing duplicated citations and using the selection criteria. We removed 5 articles^50, 63, 71, 75, 88^ from the meta-analysis for reasons shown in the ESF 1 Table 5. Consequently, 59 articles^17, 18, 20, 37-49, 51-62, 64-70, 72-74, 76-87, 89-97^ were considered in the meta-analysis (**Figure 1**).

**Figure 1.**
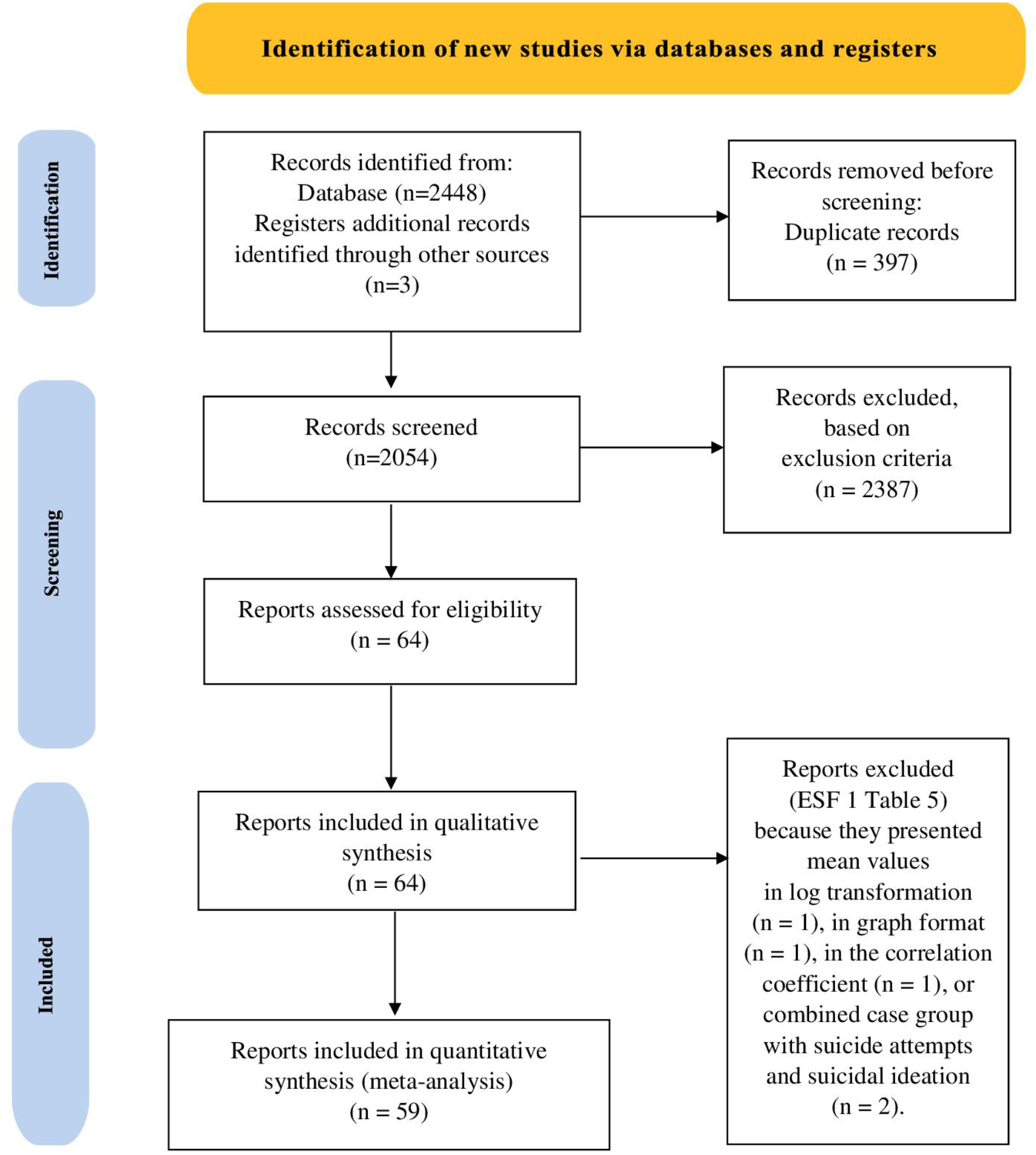
PRISMA 2020 diagram.

**ESF 1 Table 4** shows the characteristics of the included studies. Fifty-two articles were case control studies, and four articles were cohort studies, two were retrospective, and one was a longitudinal study. Thirty-three studies assayed the biomarkers in serum, 18 in plasma, 4 in blood, and 4 reported mixed mediums. Of these, only 35 studies samples reported the specific time for blood sampling. All of the study participants ages range from 12 to 89 years old. We included participants from all continents except Australasia, namely 1 study from Belgium, Croatia, Egypt, Iran, Ireland, Japan, Netherland, and Tunisia; 2 studies from Canada, France, India, Italy, Mexico, Poland, and Taiwan; 3 studies from Brazil and Iraq; 4 studies from Sweden; 6 studies from China and Turkey; 7 studies from USA; and 8 studies from South Korea. The median quality scores were 5.7 (min = 1.6, max = 9.7), whereas the median red point score was 17.5 (min = 9.0, max = 25.0).

The meta-analysis included 16.411 participants samples, namely 4.034 cases (either SA+ or SI+) and 12.377 controls. We used subgroups within the study as unit of analysis to compare 2.511 SA+ cases *versus* 7.784 SA- and heathy controls combined, and 1.523 SI+ *versus* 4.593 SI- and healthy controls combined (**Table 1**).

We found that 42.05% of the SA patients in the 59 studies belonged to a mixed group of psychiatric disorders (MIX), whereas 34.41% suffered from MDD, 19.59% of affective disorders, and 1% showed other psychiatric disorders including schizophrenia and adjustment disorders (OTHER). We found that 23.7% of the SI+ cases suffered from MDD and 23.57% from affective disorders, whilst 23.05% and 9.26% of the cases were allocated to the MIX or OTHER class. Most SA+ cases were included based on the attempted suicidal act (67.98%) and the others were included based on a semi-structured interview (32.02%), whereas most of SI+ cases were assessed by a self-report questionnaire (63.23%) and the remaining using a semi-structured interview (36.76%).

### Qualitative synthesis

Table 1 shows the distribution of the SMD and confidence intervals of the outcome profiles. Inspecting the confidence intervals showed that most included studies favored SB and that only 1 or 2 studies reporting on IO&NS, IRS, inflammation, neurotoxicity, and ANTIOXPRO showed confidence intervals which were entirely located on the negative side of zero. All studies on CRP and BDNF favored SB and no studies showed a CI, which was entirely on the negative side of zero. Furthermore, there were two studies, not included in the CMA (ESF 1 Table 5), which reported no significant findings on plasma cytokines^63^, chemokines^63^, and TRP^88^, whereas the other three studies revealed inconsistent results on plasma CRP^50, 63, 75^. Regarding the SB subtypes SA and SI, we found that most studies favor SA and that no CI was entirely located on the negative side of zero (except the ANTIOXPRO profile), whereas in the SI studies 1 or 2 confidence intervals were entirely located on the negative side of zero. In addition, there were no significant findings on the plasma TRP/amino acid ratio^71^ in studies which were not included in the CMA (ESF 1 Table 5) and which compared SA+ cases with SA-controls.

### Meta-analysis of the primary outcome: IO&NS profile

**Table 2** and **Figure 2** show that there is a significant difference in the overall pooled effect size (0.299) when comparing IO&NS in SB with controls. We used subgroup analysis within study as the unit of analysis and found that the comparison of SA+ *versus* controls yielded a SMD of 0.387, and SI+ *versus* controls a SMD of 0.167. Using subgroups as the unit of analysis, we observed that the comparison of SA+ with SA-resulted in an SMD of 0.255, but the comparison of SA+ *versus* HC resulted in a large, pooled effect size (0.796). The comparison of SI+ *versus* SI-yielded a significantly but small effect size (n=21; SMD: 0.156; 95%CI: 0.016; 0.296), whereas the SMD was not significantly different when comparing SI+ with HC. **ESF 2 Figure 1** showed the differences in the effect sizes of the comparisons of SA+ vs controls (SMD=0.387) and SI+ *versus* controls (0.167) which was significant (χ^2^=5.14, df=1, p=0.023), indicating that SA+ is more strongly associated with IO&NS than SI+. The results of these meta-analyses were unaltered by sensitivity analysis employing the leave-one-out method (and any of the meta-analyses described below). Consequently, we performed a sensitivity analysis on the high-quality studies (quality score ≥7.0; redpoint score ≤ 18.0) and compared those with lower quality studies (quality score ≤4.0; redpoint score >18.0). **ESF 2 Figure 2** shows the meta-analysis performed on the 13 high-quality studies^44, 46, 52, 53, 55, 64, 66, 69, 70, 72, 84, 91, 93^ (SMD: 0.407; 95%CI: 0.203; 0.611; τ^2^=0.092) and 5 low-quality studies^39, 73, 79, 89, 92^ (SMD: 0.129; 95%CI: 0.014; 0.0.243; τ^2^=0.005) when comparing SB to controls. This difference between high- and low-quality studies was significant (χ^2^=5.444, df=1, p=0.020).

**Table 2.** Results of meta-analysis performed on different outcome variables (immune and oxidative and nitrosative stress profiles, IO&NS, and subdomains)

| Outcome feature sets | n studies | Groups, subgroups | SMD | 95% CI | z | p | Q | df | p | I<sup>2</sup> (%) | $\tau^2$ | T |
| --- | --- | --- | --- | --- | --- | --- | --- | --- | --- | --- | --- | --- |
| IO&NS | 59 | Overall | 0.299 | 0.200; 0.397 | 5.933 | <0.001 | 276.474 | 58 | <0.001 | 79.022 | 0.100 | 0.316 |
|  | 37 | SA+ vs. Controls | 0.387 | 0.256; 0.519 | 5.771 | <0.001 | 176.200 | 36 | <0.001 | 79.569 | 0.114 | 0.337 |
|  | 21 | SA+ vs. HC | 0.796 | 0.503; 1.089 | 5.322 | <0.001 | 152.636 | 20 | <0.001 | 86.897 | 0.393 | 0.627 |
|  | 29 | SA+ vs. SA- | 0.255 | 0.140; 0.370 | 4.344 | <0.001 | 90.748 | 28 | <0.001 | 69.145 | 0.056 | 0.238 |
|  | 24 | SI+ vs. Controls | 0.167 | 0.030; 0.304 | 2.396 | 0.017 | 83.446 | 23 | <0.001 | 72.437 | 0.071 | 0.267 |
| IRS | 33 | Overall | 0.460 | 0.314; 0.605 | 6.186 | <0.001 | 174.008 | 32 | <0.001 | 81.610 | 0.128 | 0.358 |
|  | 23 | SA+ vs. Controls | 0.590 | 0.400; 0.779 | 6.101 | <0.001 | 133.065 | 22 | <0.001 | 83.467 | 0.158 | 0.398 |
|  | 14 | SA+ vs. HC | 1.049 | 0.637; 1.462 | 4.983 | <0.001 | 118.960 | 13 | <0.001 | 89.072 | 0.534 | 0.731 |
|  | 17 | SA+ vs. SA- | 0.384 | 0.229; 0.538 | 4.855 | <0.001 | 56.249 | 16 | <0.001 | 71.555 | 0.064 | 0.254 |
|  | 11 | SI+ vs. Controls | 0.231 | 0.033; 0.430 | 2.281 | 0.023 | 28.265 | 10 | 0.002 | 64.621 | 0.063 | 0.251 |
| Inflammation | 25 | Overall | 0.459 | 0.284; 0.633 | 5.142 | <0.001 | 147.659 | 24 | <0.001 | 83.746 | 0.146 | 0.382 |
|  | 16 | SA+ vs. Controls | 0.623 | 0.388; 0.858 | 5.199 | <0.001 | 95.325 | 15 | <0.001 | 84.264 | 0.173 | 0.415 |
|  | 9 | SA+ vs. HC | 1.200 | 0.584; 1.816 | 3.820 | <0.001 | 90.393 | 8 | <0.001 | 91.150 | 0.791 | 0.889 |
|  | 13 | SA+ vs. SA- | 0.414 | 0.223; 0.605 | 4.250 | <0.001 | 47.029 | 12 | <0.001 | 74.484 | 0.080 | 0.282 |
|  | 10 | SI+ vs. Controls | 0.239 | 0.000; 0.478 | 1.962 | 0.050 | 36.317 | 9 | <0.001 | 75.218 | 0.099 | 0.315 |
| CRP | 15 | Overall | 0.573 | 0.366; 0.780 | 5.433 | <0.001 | 99.036 | 14 | <0.001 | 85.864 | 0.128 | 0.358 |
|  | 10 | SA+ vs. Controls | 0.689 | 0.386; 0.991 | 4.463 | <0.001 | 78.523 | 9 | <0.001 | 88.538 | 0.194 | 0.440 |
|  | 4 | SA+ vs. HC | 1.311 | 0.446; 2.177 | 2.969 | 0.003 | 32.712 | 3 | <0.001 | 90.829 | 0.704 | 0.839 |
|  | 8 | SA+ vs. SA- | 0.499 | 0.234; 0.764 | 3.696 | <0.001 | 43.082 | 7 | <0.001 | 83.752 | 0.110 | 0.331 |
|  | 6 | SI+ vs. Controls | 0.429 | 0.161; 0.697 | 3.138 | 0.002 | 18.524 | 5 | 0.002 | 73.009 | 0.075 | 0.274 |
| Neurotoxicity | 29 | Overall | 0.403 | 0.253; 0.553 | 5.262 | <0.001 | 133.901 | 28 | <0.001 | 79.089 | 0.116 | 0.340 |
|  | 19 | SA+ vs. Controls | 0.524 | 0.320; 0.727 | 5.048 | <0.001 | 94.676 | 18 | <0.001 | 80.988 | 0.147 | 0.383 |
|  | 12 | SA+ vs. HC | 0.904 | 0.431; 1.378 | 3.745 | <0.001 | 106.480 | 11 | <0.001 | 89.669 | 0.607 | 0.779 |
|  | 14 | SA+ vs. SA- | 0.403 | 0.218; 0.588 | 4.278 | <0.001 | 49.022 | 13 | <0.001 | 73.481 | 0.079 | 0.282 |
|  | 11 | SI+ vs. Controls | 0.243 | 0.036; 0.449 | 2.299 | 0.021 | 30.692 | 10 | 0.001 | 67.418 | 0.072 | 0.268 |
| ANTIOXPRO | 40 | Overall | 0.179 | 0.063; 0.294 | 3.031 | 0.002 | 196.268 | 39 | <0.001 | 80.129 | 0.095 | 0.308 |
|  | 26 | SA+ vs. Controls | 0.199 | 0.058; 0.341 | 2.761 | 0.006 | 119.840 | 25 | <0.001 | 79.139 | 0.091 | 0.302 |
|  | 13 | SA+ vs. HC | 0.476 | 0.083; 0.869 | 2.372 | 0.018 | 118.374 | 12 | <0.001 | 89.863 | 0.461 | 0.679 |
|  | 22 | SA+ vs. SA- | 0.137 | 0.021; 0.253 | 2.322 | 0.020 | 58.340 | 21 | <0.001 | 64.004 | 0.039 | 1.197 |
|  | 16 | SI+ vs. Controls | 0.141 | -0.056; 0.338 | 1.401 | 0.161 | 73.159 | 15 | <0.001 | 79.497 | 0.113 | 0.336 |
| BDNF | 10 | Overall | 0.479 | 0.224; 0.735 | 3.681 | <0.001 | 34.811 | 9 | <0.001 | 74.146 | 0.118 | 0.344 |
|  | 7 | SA+ vs. Controls | 0.417 | 0.102; 0.733 | 2.590 | 0.010 | 25.883 | 6 | <0.001 | 76.819 | 0.131 | 0.362 |
|  | 4 | SA+ vs. HC | 0.962 | 0.708; 1.217 | 7.416 | <0.001 | 2.628 | 3 | 0.453 | 0.000 | 0.000 | 0.000 |
|  | 6 | SA+ vs. SA- | 0.256 | -0.026; 0.538 | 1.779 | 0.075 | 14.800 | 5 | 0.011 | 66.217 | 0.076 | 0.275 |
|  | 3 | SI+ vs. Controls | 0.665 | 0.413; 0.917 | 5.169 | <0.001 | 2.039 | 2 | 0.361 | 1.934 | 0.001 | 0.032 |

SMD: standardized mean difference, 95% CI: 95% confidence intervals
IO&NS: immune-inflammatory and oxidative and nitrosative stress
IRS: immune-inflammatory response system
ANTIOXPRO: protection via antioxidants and neurotrophic products.
CRP: C-reactive protein
BDNF: Brain-derived neurotrophic factor
We used the study as well as the prespecified subgroups as the units of analysis. Thus, patients with suicide behaviors (SB) were first (overall) compared with controls. Second, we performed subgroup analyses comparing the pooled effect size of IO&NS, IRS, inflammation, CRP, neurotoxicity, ANTIOXPRO and BDNF profiles in a) SA+ *versus* SA- patients or healthy controls); and b) SI+ *versus* SI- patients or healthy controls).

**Figure 2.**
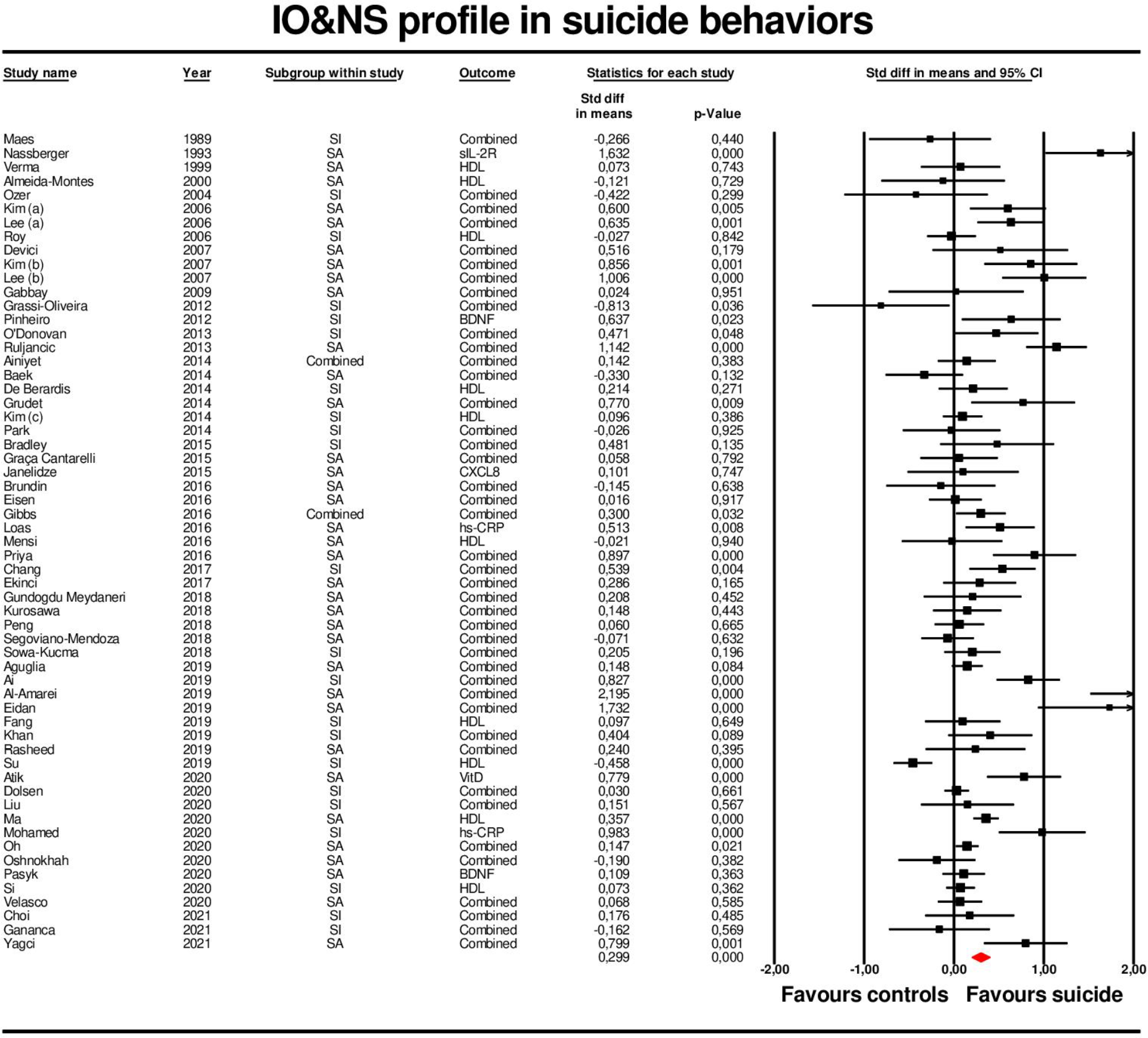
Forest plot with results of meta-analysis performed on 59 studies reporting immune-inflammatory and oxidative & nitrosative stress (IO&NS) biomarkers.

### Meta-analyses of the secondary outcomes

#### IRS profile

Table 2 reports a moderate, pooled effect size of 0.460 in overall SB cases. Subgroup analysis showed that the comparison of SA+ *versus* controls yielded a significantly effect size of 0.590, whereas SI+ *versus* controls yielded a lower effect size of 0.231. When using subgroups as the unit of analysis, we found that the comparison of SA+ *versus* SA-yielded a SMD of 0.384, whereas SA+ *versus* HC yielded a large, pooled effect size of 1.049. There was no significant difference between SI+ and SI- or HC groups. The difference in the effect sizes of the comparisons of SA+ vs controls (SMD=0.590) and SI+ *versus* controls (0.231) was significant (χ^2^=6.54, df=1, p=0.011).

#### Inflammatory profile

We found that the inflammation score is higher in SB cases than in controls with a moderate effect size of 0.459. Subgroup analysis showed that the comparison of SA+ *versus* controls yielded a SMD of 0.623, whereas the comparison of SI+ *versus* controls yielded a SMD of 0.239 (**Figure 3**). When using the subgroups as the unit of analysis, we found that the comparison of SA+ *versus* SA-yielded a SMD of 0.414, whereas SA+ *versus* HC yielded a large, pooled effect size of 1.200. There was no significant difference between SI+ and SI- or HC groups. The difference in the effect sizes of the comparisons of SA+ vs controls (SMD=0.623) and SI+ *versus* controls (SMD=0.239) was significant (χ^2^=5.048, df=1, p=0.025).

**Figure 3.**
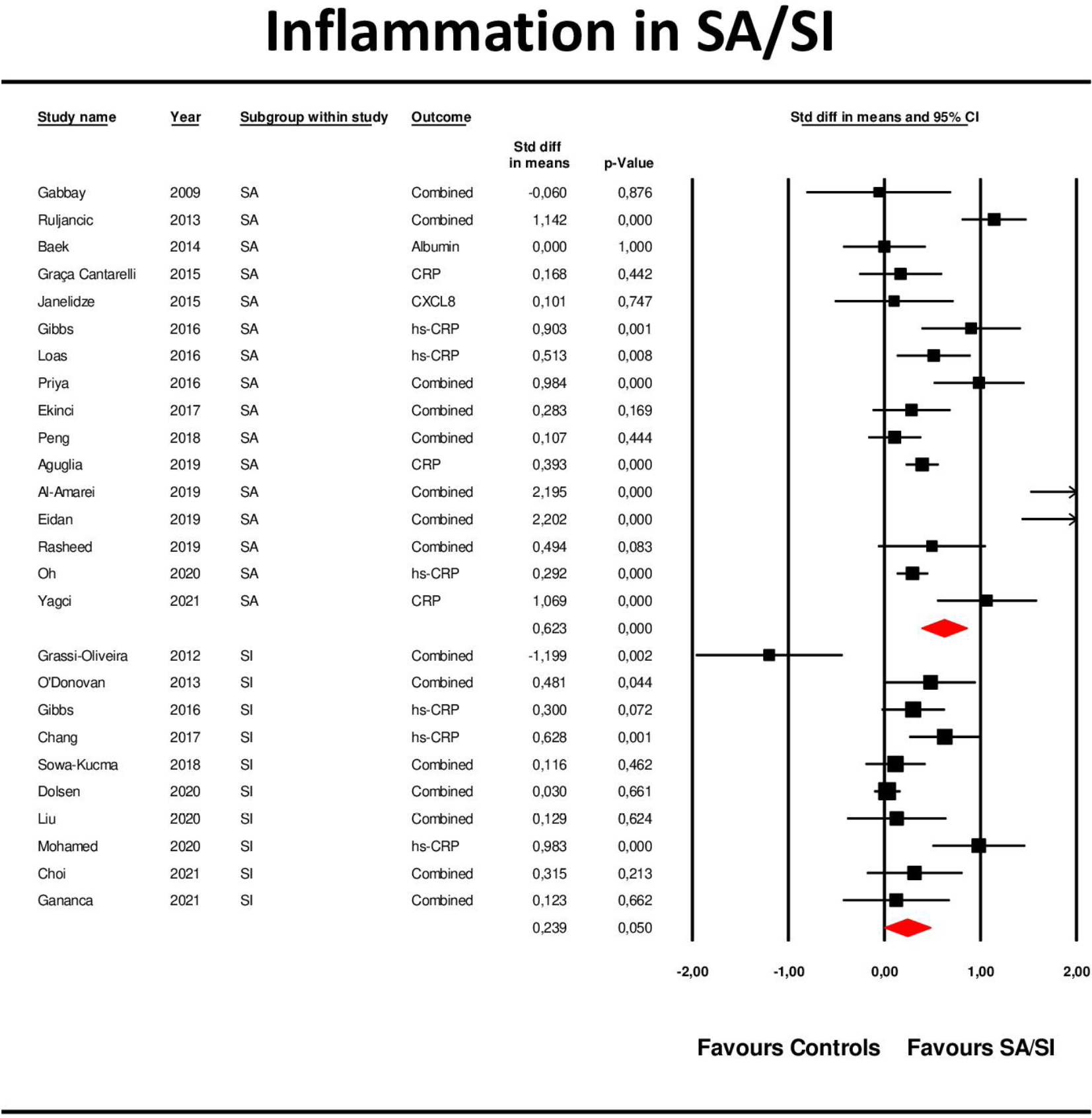
Forest plot with results of meta-analysis (subgroup analysis) performed on 26 studies reporting inflammation profile.

#### CRP

When comparing CRP in SB with controls, we found a pooled effect size of 0.573 (table 2). Subgroup analysis showed that the comparison of SA+ *versus* controls yielded a significant pooled effect size of 0.689, whereas SI+ *versus* controls yielded a significant effect size of 0.429. When using subgroups as the unit of analysis, we found that the comparison of SA+ *versus* SA-yielded a SMD of 0.499, whereas SA+ *versus* HC yielded a large, pooled effect size of 1.311. The comparison of SI+ *versus* SI-yielded a significantly moderate effect size (n=4; SMD: 0.460; 95%CI: 0.098; 0.822). The comparison of SI+ *versus* HC showed only two studies and therefore we did not perform CMA. The difference in the effect sizes of the comparisons of SA+ vs controls and SI+ *versus* controls was not significant (χ^2^=1.58, df=1, p=0.208).

#### Neurotoxicity profiles

Table 2 also shows that the neurotoxicity levels are significantly increased in SB as compared with controls with an overall pooled effect size of 0.403. Subgroup analysis (with the subgroup as the unit of analysis) showed that the comparison of SA+ *versus* controls yielded a significant pooled effect size of 0.524, whereas SI+ *versus* controls yielded a lower effect size of 0.243. We found that the comparison of SA+ *versus* SA-yielded a SMD of 0.403, whereas SA+ *versus* HC yielded a large, pooled effect size of 0.904. There was no significant difference between SI+ and SI- or HC groups. The difference in the effect sizes of the comparisons of SA+ vs controls and SI+ *versus* controls was not significant (χ^2^=3.61, df=1, p=0.057).

#### ANTIOXPRO profiles

We found significant associations between ANTIOXPRO scores in SB cases with a small, pooled effect size of 0.179. The comparison of SA+ *versus* controls yielded a low but significant effect size of 0.199, whereas there were no significant differences when comparing SI+ and controls. When using the subgroups as the unit of analysis, we found that the comparison of SA+ *versus* SA-yielded a SMD of 0.137, whereas SA+ *versus* HC yielded a SMD of 0.476. The difference in the effect sizes of the comparisons of SA+ vs controls and SI+ *versus* controls was not significant (χ^2^=0.22, df=1, p=0.636).

#### BDNF

Table 2 shows that SB is accompanied by the lowered BDNF levels with a SMD of 0.479. The comparison of SA+ *versus* controls yielded a SMD of 0.417, and SI+ *versus* controls yielded a SMD of 0.665. When using the subgroups as the unit of analysis, we found that the comparison of SA+ *versus* SA-yielded a SMD of 0.256, whereas SA+ *versus* HC yielded a large, pooled effect size of 0.962. The comparison of SI+ *versus* SI-yielded a significantly moderate effect size (n=3; SMD: 0.587; 95%CI: 0.324; 0.851). We are unable to perform the analyses of SI+ *versus* HC due to the small number of studies (n=2). The difference in the effect sizes of the comparisons of SA+ vs controls and SI+ *versus* controls was not significant (χ^2^=1.44, df=1, p=0.230).

### Other subgroup analyses and meta-regression analyses

**ESF 2 Table 1** shows the association among IO&NS profiles and SB remained significant in patients with MDD and MIX, whereas in patients belonging to the affective and OTHER groups they were not significant. In addition, these inter-class differences were significant (χ^2^=8.563; df=3; p=0.036). The IO&NS markers showed a significant association with SB in inpatients but not in outpatients. The comparisons of in- and outpatients in SB *versus* controls (χ^2^ = 8.147; df=1; p=0.004) and SA *versus* controls (χ^2^ = 5.770; df=1; p=0.016) were significantly different. There were no significant differences in effect sizes obtained in studies using serum, plasma, and blood (χ^2^ =4.677; df=2; p=0.096). There are significant associations between IO&NS markers and SB in patients who are diagnosed based on the suicidal act or an interview, but not when the SB was assessed through self-report, and this difference was significant (χ2 = 8.479; df=2; p=0.014). In SA, the IO&NS effect size was significantly larger in studies which diagnosed SB by registering the act as compared with studies which used an interview (χ^2^ =7.102; df=1; p=0.008). In SI, there was no significant difference between interview and self-report (χ^2^ = 0.015; df=1; p=0.902). Meta-regression revealed no effects of latitude, quality scores, and redpoint scores.

### Publication bias

**ESF 2 Table 2** shows the impact of publication bias on the studies under the random-effect model. There is some degree of publication bias for IO&NS, IRS, inflammation, CRP, neurotoxicity, and ANTIOXPRO studies with 6, 5, 3, 2, 4, and 4 missing studies samples on the right side of the funnel plot, respectively, and correction yielded an increase in the overall adjusted point estimate. There was no impact of publication bias on the BDNF studies.

## Discussion

The first major finding of this systematic review and meta-analysis is that SB (namely SA and SI) is accompanied by significantly increased levels of the primary outcome variable, the IO&NS profile, albeit with a small effect size. In addition, also the IRS, inflammation (including CRP), neurotoxicity, ANTIOXPRO (including BDNF) profiles were significantly associated with SB, with small to moderate effect sizes. Our meta-analysis thus confirms that the pathophysiology of SB is related to activated IO&NS pathways ^4, 22, 98^ with elevated CRP^26^ and reduced BDNF levels.^28^ Most importantly, our results show that suicidal behaviors are associated with an inflammatory response which is mainly driven by M1 macrophage-derived cytokines^11, 99^ including IL-1β, IL-6, and TNF-α, which induce the production of acute phase proteins such as CRP and downregulate albumin, and by Th-1 cells with increased production of IFN-γ^11^. These products of activated macrophages and T cells exert neurotoxic effects thereby affecting white and gray matter plasticity^11^. Furthermore, both M1 and Th-1 cytokines may stimulate indoleamine 2,3-dioxygenase (IDO), lowering plasma TRP and inhibiting 5-HT synthesis, as well as increasing the production of neurotoxic TRYCATs such as KYN and QA, which may result in N-Methyl-D-Aspartic acid (NMDA)-induced neurotoxicity^8, 99^. Furthermore, increased indicants of nitro-oxidative stress with increased lipid peroxidation and lowered antioxidant levels, further contribute to increased neurotoxicity and reduced neuroprotection.^7^ Chronically elevated IO&NS pathways cause breakdown of the blood brain barrier (BBB), which allows neurotoxic molecules to enter the brain thereby contributing to the neurotoxic effects. Previously, it was shown that activated IO&NS pathways are associated with major depressive and bipolar disorder, staging of affective disorders, neurodegenerative processes, and schizophrenia.^11, 99, 100^

We should stress that the present meta-analysis study used composite scores reflecting IO&NS profiles.^11, 12, 101^ Previously, we have discussed the many advantages of using composite scores instead of solitary markers in case-control studies ^11, 12, 101^ and in meta-analysis as well.^18^ For example, if a study reports on two inflammatory markers (e.g. CRP and albumin), one of which is significantly associated with SA but the other is not, using only one biomarker in a meta-analysis may result in incorrect conclusions about “inflammation”^19^. Therefore, performing meta-analyses on one biomarker only may not allow generalizing the findings because the results may by quite different after including all biomarkers of the same profile. Moreover, the selection of only one biomarker per study may induce more biological between-study variance because different confounding variables may differently affect the biomarkers of the same profile.^19^ For example, selecting CRP as an index of inflammation is prone to bias because CRP is affected by many confounding variables including early lifetime trauma, sex, age and especially body mass index^19, 102, 103^. In our study, the different biomarker scores were not only averaged in one and the same study but also averaged over all studies. As such, our meta-analyses used more robust synthetic indices of IO&NS and its subdomains based on all available data and, therefore, these data allow to generalize the findings to the functional profiles specified in our study.

The second major finding of this study is that the effects sizes of the IO&NS, IRS, inflammation, CRP, and neurotoxicity scores were higher in SA than in SI, and that the effect sizes of IO&NS, IRS and inflammation were significantly greater in SA than in SI. These findings may indicate that IRS and inflammatory processes are more tightly associated with SA than with SI. This may point towards differences in the severity with SA being more severe than SI^1^. On the other hand, the effects sizes of the comparisons between SA and SI were not significant when considering CRP, neurotoxicity, ANTIOXPRO and BDNF. Therefore, the results of the current meta-analysis do not confirm the results of Gibbs et al.^57^ who reported significant differences in serum hs-CRP levels between SA and SI. All in all, these results show that the higher IO&NS and IRS profiles in SA *versus* SI should be attributed to smaller but cumulative effects of inflammation, neurotoxicity, and lowered neuroprotection in SA.

The study showed some statistical heterogeneity as assessed using the tau (τ) statistic, which represents a precise measurement of heterogeneity and which is insensitive to the number of patients or studies in the meta-analysis.^35, 36^ On the other hand, Q-values and the *I*^*2*^ metric do not reflect the degree of heterogeneity, although frequently employed as such.^18^ Nevertheless, the current meta-analysis was able to detect five possible sources of statistical heterogeneity. Firstly, differences in outpatient *versus* inpatient status with higher effect sizes in the latter. This may be explained by effects of severity of illness which is higher in inpatients than outpatients^104^, and the higher frequency of SA in inpatients^104^. The second source of heterogeneity concerns the assessment of SB with a greater effect size when the diagnosis of SA was based on the suicidal act rather than on an interview. These results show that the assessment of SB based on interview and self-report may largely underestimate the actual effect sizes and, therefore, are inaccurate assessment methods. Future research on SB should recruit patients with SA based on the registered suicidal act.

Thirdly, the quality of the included papers contributed to heterogeneity as evidenced by higher pooled effect sizes of IO&NS in high-quality studies than low-quality studies. The fourth source of statistical heterogeneity is due to the choice of controls, namely patients without SB *versus* heathy controls. As such, we found that that the pooled effect sizes of the IO&NS, IRS, inflammation (including), CRP, and neurotoxicity profiles were larger when comparing SB with healthy controls as reference group (between 0.796 and 1.311), whereas lower effect sizes were obtained when comparing patients with and without SB (between 0.3 to 0.5). The comparisons with normal controls show that activated IO&NS pathways are strongly associated with SA in patients with psychiatric disorders, and the comparisons obtained with patients without SB show that SB is accompanied by aberrations in IO&NS pathways above and beyond the increased levels in psychiatric patients. Therefore, it is safe to conclude that activated IO&NS, IRS, inflammatory and neurotoxic pathways and lowered neuroprotection are strongly associated not only with the major psychiatric disorders ^11, 101^ but also with the SB which are associated with those disorders.

Fifthly, inspection of the forest plots shows that the heterogeneity is increased by a number of heterogeneity-influencing studies which favor SA. Previously, we performed sensitivity analyses on the included studies after deselecting a few heterogeneity-influencing studies and found that the prediction intervals no longer overlapped with the zero SMD but fell completely on the right site of zero. Indeed, a few small n studies that favor SA and show a very large effect size may bias SMD whereby the heterogeneity may increase in a random effect model and the latter may be more biased that when using a fixed effect model^19^. It should be added that there is probably nothing wrong with these heterogeneity-influencing studies, they only show a large SMD.

Several limitations should be considered when discussing our findings. Firstly, there are very few studies which assayed IL-4^56, 72^ and TGF-β1^78^ and therefore these important Th-2 and T regulatory (Treg) cytokines could not be included in the CMA. Secondly, key biomarkers of other profiles were also missing. For example, haptoglobin, transferrin, and zinc were missing as inflammatory biomarkers (and its parent profiles). Vitamin E, coenzyme Q10, glutathione, catalase, MPO and xanthine oxidase were missing as O&NS biomarkers. In fact, we were even unable to compute a CIRS profile, although most important to evaluate the homeostatic processes in the immune system.^11^ As a result, future research should concentrate on the missing biomarkers, such as Treg cytokines like IL-10 and TGF-β1, as well as Th-2 cytokines like IL-4, IL-5 and IL-13 (see also ESF 1 Table 1; column “what is missing in SB research”). Thirdly, this study did not include other potential variables regarding severity of SB, including lifetime SB^19^, staging of the major psychiatric disorders^13^, and stressful live events, including early lifetime trauma^4^. Further studies should also include those covariates.

In conclusion, this is the first meta-analysis to explore IO&NS pathways in SA *versus* SI. Recent SB (within 3 months) is accompanied by aberrations in IO&NS profiles, which are more strongly associated with SA than SI. Increased neurotoxicity due to inflammation and nitro-oxidative stress may explain why psychiatric patients are at higher risk of SA and SI than healthy controls. As such, we have delineated new drug targets to treat SB, namely the IO&NS subdomains.

## Supporting information

ESF 1

ESF 2

## Data Availability

The dataset (CMA file) generated during and/or analyzed during the current study will be available from MM upon reasonable request and once the dataset has been fully exploited by the authors.

## Ethical approval and consent to participate

Not applicable.

## Consent for publication

Not applicable.

## Author’s contributions

All authors contributed to the writing up of the paper. The work was designed by MM and AV. Data were collected by AV and KJ. Statistical analyses were performed by MM and AV. All authors revised and approved the final draft.

## Acknowledgements

Not applicable.

## Compliance with Ethical Standards/Disclosure of potential conflicts of interest

The authors have no conflicts of interest to declare that are relevant to the content of this article.

## Research involving Human Participants and/or Animals

Not applicable.

## Informed consent

Not applicable.

