## Supplementary material for "Inflammation and nitro-oxidative stress in current suicidal attempts and current suicidal ideation: a systematic review and meta-analysis": ESF 1

**Electronic Supplementary Information File (ESF) 1**

Activated immune, oxidative and nitrosative stress pathways are strongly associated with suicidal attempts and less with suicidal ideation: a meta-analysis and meta-regression.

**Running title:** immune activation in suicidal behaviors

Asara Vasupanrajit, M.Sc.^a^; Ketsupar Jirakran, M.Sc.^a,b^; Chavit Tunvirachaisakul, M.D., Ph.D.^a,^ ^c^; Michael Maes, M.D., Ph.D.^a, c, d, e^

^a^ Department of Psychiatry, Faculty of Medicine, Chulalongkorn University, Bangkok, Thailand.

^b^ Maximizing Thai Children’s Developmental Potential Research Unit, Department of Pediatrics, Faculty of Medicine, Chulalongkorn University, Bangkok, Thailand.

^c^ Cognitive Impairment and Dementia Research Unit, Department of Psychiatry, Faculty of Medicine, Chulalongkorn University, Bangkok, Thailand.

^d^ IMPACT Strategies Research Center, Deakin University, Geelong, Australia.

^e^ Department of Psychiatry, Medical University of Plovdiv, Plovdiv, Bulgaria.

**ESF 1 Table 1.** Explanation of peripheral immune and O&NS phenotypes.

| Immune and O&NS phenotype | | What is | What examined | What is missing in SA research |
| --- | --- | --- | --- | --- |
| IRS^1, 2^ | Inflammation (M1) | M1 cytokines, Acute Phase Proteins (APR), Complement | Interleukin (IL)-6, Tumor necrosis factor (TNF)-α,  IL-1β,  C-reactive protein (CRP), Albumin, CCL2, CXCL8 | Fibrinogen sIL-6R, soluble gp130, sTNF-αR1, sIL-1RA, complement factors |
|  | T helper (Th)-1 | IL-2, Interferon (IFN)-γ, IL-12 | IL-2, IFN-γ, IL2/IL4, IFN-γ/IL4 | IL-12 |
|  | Cell-mediated immunity (CMI) | Interaction M1 and Th1 | IL-6, TNF-α, IL-1, IL-2, IFN-γ,  sIL-2R, TRYCATS (i.e., PA, QA, KYN), Tryptophan | Neopterin |
|  | Th-17 | IL-6, IL-17 | IL-6 | IL-17 |
|  | Chemokines | CXCL-8, eotaxin | CXCL-8, CCL2, CCL11, RANTES | eotaxin, etc |
|  | APR proteins | Plasminogen Activator Inhibitor-1 (PAI-1), Haptoglobin (Hp), serum amyloid A and P, alpha-1 acid, complement system  (such as C3 and C4), CRP, Fb, Albumin | CRP, Albumin | Fibrinogen, Hp,  PAI-1, retinol binding protein (RBP) |
| CIRS^1, 2^ | Regulatory T cells  (T-reg) | IL-10, Transforming growth factor (TGF)-β | TGF-β | T-reg cells  CD4, CD25, Forkhead box P3 (FOXP3), TGF-β |
|  | Th-2 | IL-4, IL-5, IL-9, IL-10, IL-13 and IL-25 | IL-4 | IL-13, IL-5, IL-9 |
| O&NS^1, 2^ | Nitro-oxidative stress | Advanced oxidation protein products (AOPP), Homocysteine, Lipid Hydroperoxide (LOOH), 3,4-Methylenedioxyamphetamine (MDA), Nitric oxide metabolites (NOx) | MDA, NOx, TBARS | DNA oxidation damage to mitochondria, AOPP, Homocysteine, LOOH |
|  | Antioxidant | Albumin, BDNF, HDL-c, total reactive antioxidant potential (TRAP), Vitamin D | Albumin, BDNF, DHA, EPA, HDL-c, TAC, Tryptophan, Vitamin D | Catalase,  Vitamin A, Vitamin C, Vitamin E, Coenzyme Q10, Zinc, Glutathione |
| NT | Neurotoxicity^3^ | IRS, CIRS, Nitro-oxidative stress | AA, CCL1, CCL2, CCL3, RANTES, CCL11, CCL17, CCL22, CCL23, CCL24, CCL27, CX3CL1, CXCL2, CXCL5, CXCL8, CXCL10, CXCL12, CXCL16, CRP, IFN-γ, IL-1β, IL-2, IL-6, TNF-α, TRYCATS (i.e., PA, QA, KYN), MDA, NOx, TBARS, TNFR60, TNFR80 | LPS bacteria, indicants BBB, AOPP, Homocysteine, LOOH, |
| NP | Neuroprotection^3^ | Antioxidant | Albumin, BDNF, DHA, EPA, HDL-c, TAC, Trytophan, Vitamin D | Vitamin E, Coenzyme Q10, Zinc, Glutathione |

Note: Further explanation of the function of the peripheral immune and O&NS phenotypes can be found in Maes and Carvalho, 2018^1^; Roomruangwong et al., 2020^2^ and Maes et al., 2021^3^

1. Maes M, Carvalho AF. The Compensatory Immune-Regulatory Reflex System (CIRS) in Depression and Bipolar Disorder. Mol Neurobiol 2018; 55(12): 8885-8903.
2. Roomruangwong C, Noto C, Kanchanatawan B, Anderson G, Kubera M, Carvalho AF et al. The Role of Aberrations in the Immune-Inflammatory Response System (IRS) and the Compensatory Immune-Regulatory Reflex System (CIRS) in Different Phenotypes of Schizophrenia: the IRS-CIRS Theory of Schizophrenia. Mol Neurobiol 2020; 57(2): 778-797.
3. Maes M, Moraes JB, Bonifacio KL, Barbosa DS, Vargas HO, Michelin AP et al. Towards a new model and classification of mood disorders based on risk resilience, neuro-affective toxicity, staging, and phenome features using the nomothetic network psychiatry approach. Metabolic brain disease 2021; 36(3): 509-521.

Abbreviations:

AA: Arachidonic acid (omega-6 fatty acid)

APR: the acute phase response

BBB: Blood-Brain Barrier

BDNF: Brain-derived neurotrophic factor

CCL: The C–C motif chemokine ligand

CIRS: The compensatory immune-regulatory reflex system

CXCL: The chemokine C-X-C motif ligand

CX3CL1: The C-X3-C Motif Chemokine Ligand 1/Fractalkine

DHA: Docosahexaenoic acid (omega-3 fatty acid)

EPA: Eicosapentaenoic acid (omega-3 fatty acid)

HDL-c: High-density lipoprotein cholesterol

IRS: The immune-inflammatory response system

KYN: Kynurenine

LPS: Lipopolysaccharides

MDA: Malondialdehyde

NOx: Nitric oxide metabolites

NT: Neurotoxicity

NP: Neuroprotection

O&NS: Antioxidants and nitro-oxidative stress

PA: Picolinic acid

QA: Quinolinic acid

RANTES: Regulated on Activation, Normal T Cell Expressed and Secreted

SA: Suicide attempts

SI: Suicide ideation

sIL2R: Soluble Interleukin 2 Receptor

sIL6R: Soluble Interleukin 6 Receptor

sTNF-αR1: Soluble tumor necrosis factor alpha receptor 1

TAC: Total antioxidant capacity

TBARS: Thiobarbituric acid reactive substances

TNFR: Tumor necrosis factor receptors

TRYCATs: The tryptophan catabolites

**ESF 1 Table 2.** Specific search for each database.

**PubMed/Medline**

((inflamm* OR immun*) AND (cytokine OR chemokine OR il-6 OR il-1 OR interleukin OR (c-reactive protein OR CRP) OR (Tumor necrosis factor OR tnf) OR (interferon OR ifn) OR (Transforming growth factor OR tgf) OR Tryptophan AND (suicid*)) **1072**

**PubMed/Medline**

((Oxidative stress OR Antioxidants) AND (Zinc) OR (Vitamin C) OR (Albumin) OR (Nitric oxide metabolites) OR (Lipid hydroperoxides) OR (Omega 3) OR (Coenzyme Q10) OR (DHA) AND (suicid*) **406**

**Google Scholar**

Suicide AND [inflamm*] OR [immun*] AND [oxidative stress biomarkers] AND antioxidants OR [cytokine] OR chemokine] OR [il-6 OR il-1 OR interleukin] OR [(c-reactive protein) OR CRP] OR [(Tumor necrosis factor) OR tnf] OR [interferon OR ifn] OR (Transforming) **617**

***WEB OF SCIENCE***

TOPIC: (Suicide AND Oxidative Stress) **353**

**^ESF 1 Table 3.^** ^Immune confounder of suicide behaviors scale (ICS); applied from Andrés-Rodríguez, et al., 2019a and Vasupanrajit et al., 2021b^

| **Methodological quality of the study** | |
| --- | --- |
| 1 | Study sample ≥ 128 participants including patients and controls (1= Yes, 0 = No) |
| 2 | Did the study control results for potential confounders (e.g., age, BMI, gender, race)? (1= Yes, 0 = No) |
| 3 | Were participants with suicide attempts (SA) and controls age- and-gender-matched or statistically controlled? (1= Yes, 0 = No) |
| 4 | Was the time of sample collection specified (e.g., morning vs. evening)? (1= Yes, 0 = No) |
| 5 | Were participants with SA free of immunomodulatory drugs including anti-cytokines, corticoids, immunoglobulins, and immunosuppressants, been through a medication washout or the intake was statistically controlled? (1= Yes, 0 = No) |
| 6 | Were participants with suicidal behaviour free of antidepressants and mood stabilizers or statistically controlled? (1= Yes, 0 = No) |
| 7 | Reporting of either the manufacturer of the test or its parameters (detection limit and coefficient of variation) (1= Yes, 0 = No) |
| 8 | Reporting how data under detection limit was handled (1 = Yes, 0 = No) |
| 9 | Reporting % of the sample under detection limit (1=Yes, 0= No) |
| 10 | Reporting blood fraction (serum, plasma, culture supernatant or whole blood) (1= Yes, 0 = No) |
|  | **Total quality score (10 points)** |
| **Biomarker confounders red points**  *The red points should not be given if the item is statistically controlled* | |
| 1 | 3 red points for comorbid illnesses such as autoimmune disorders & other immune disorders including RA, psoriasis, IBD, COPD, MS |
| 2 | 3 red points for use of recreational drugs such as methamphetamine or opioids (Not applicable if psychiatric disorders are excluded) |
| 3 | 2 red points when groups were significantly difference or there was no statistically controlled for age |
| 4 | 2 red points when groups were significantly difference or there was no statistically controlled for sex |
| 5 | 2 red points for medication use as for example immunomodulators |
| 6 | 2 red points for early traumatic life events |
| 7 | 2 red points for shift work and primary sleep disorders |
| 8 | 1.5 red points for antidepressants |
| 9 | 1 red point for more common systemic immune disorders including diabetes type 1/2, essential hypertension, metabolic syndrome |
| 10 | 1 red point for not fasting (8 hours before blood extraction) |
| 11 | 1 red point for use of omega-3 and antioxidant supplements |
| 12 | 1 red point when groups were significantly difference or there was no statistically controlled for BMI |
| 13 | 1 red point for physical activity or sedentary life |
| 14 | 1 red point for smoking |
| 15 | 1 red point for use of oral contraceptives or NSAIDs |
| 16 | 0.5 red points for ethnicity in countries such as US, Brazil (not China or Japan) |
| 17 | 0.5 red points for seasonality |
| 18 | 0.5 red points for diurnal variation (8-10 a.m. versus all other time points) |
|  | **Total red point score (26 points)** |

Note: Threshold of study samples is stablished as it is the minimum needed for statistical power. Confounders red points should be given when the item is not reported (or statistically controlled).

^a^ Andrés-Rodríguez L, Borràs X, Feliu-Soler A, Pérez-Aranda A, Angarita-Osorio N, Moreno-Peral P et al. Peripheral immune aberrations in fibromyalgia: A systematic review, meta-analysis and meta-regression. Brain, Behavior, and Immunity 2020; **87**: 881-889.

^b^ Vasupanrajit A, Jirakran K, Tunvirachaisakul C, Maes M. Suicide attempts are associated with activated immune-inflammatory, nitro-oxidative, and neurotoxic pathways: A systematic review and meta-analysis. Journal of Affective Disorders 2021; **295**: 80-92.

**ESF 1 Table 4.** Characteristics of 59 studies included in quantitative synthesis (meta-analysis).

| **Authors (Year)** | **Sub groups** | **N** | | **Psychiatric disorders in cases** | **Suicide assessment** | **Exclusion of comorbid illnesses** | **Specific time of blood sample** | **Medium** | **IO&NS markers** | **Key findings** | **Quality score**  **(max=10)** | **Redpoint score**  **(max=26)** |
| --- | --- | --- | --- | --- | --- | --- | --- | --- | --- | --- | --- | --- |
|  |  | **Cases** | **Controls** |  |  |  |  |  |  |  |  |  |
| Aguglia et al. (2019) | SA | 432 | 200 | Mixed | After SA | No | Yes | Serum | - CRP - HDL | - CRP levels in SA+ were significantly higher than in SA-, whereas no significant difference in HDL. | 6.9 | 18.5 |
| Ai et al.  (2019) | SI | 84 | 66 | MDD | Self-report | No | No | Plasma | - BDNF | - No significant difference in BDNF between SI+ and SI-. | 4.7 | 18.0 |
| Ainiyet & Rybakowski (2014) | SA, SI | 65 | 159 | Affective | Interview | Partly | Yes | Serum | - HDL | - No significant difference in HDL was observed. | 3.8 | 19.5 |
| Al-Amarei  et al. (2019) | SA | 22 | 34 | MDD | After SA | Partly | Partly | Plasma | - CRP | - CRP levels in SA+ were significantly higher than in SA- or HC. | 4.0 | 15.0 |
| Almeida-Montes  et al. (2000) | SA | 18 | 15 | MDD | After SA | Yes | Yes | Serum | - HDL | - No significant difference in HDL between SA+ and SA-. | 6.3 | 11.5 |
| Atik et al. (2020) | SA | 59 | 42 | Mixed | After SA | No | No | Serum | - Vitamin D | - Vitamin D levels in SA+ were significantly lower than in HC. | 4.1 | 20.5 |
| Baek et al. (2014) | SA | 22 | 464 | MDD | Interview | No | Yes | Serum | - Albumin - HDL | - HDL levels in SA+ were significantly higher than in SA-. - Albumin levels were not significantly different. | 5.7 | 19.0 |
| Bradley et al. (2015) | SI | 19 | 26 | MDD | Self-report | Partly | Yes | Plasma | - KYN - TRP - KYN/TRP | - TRP levels in SA+ were significantly lower than in SA- or HC, whereas KYN levels were not substantially different. - KYN/TRP ratio in SA+ were significantly higher than in SA- or HC. | 7.2 | 12.0 |
| Brundin et al. (2016) | SA | 18 | 29 | MDD, Affective | After SA | No | Yes | Plasma | - PA - QA | - PA levels in SA+ with mixed group of psychiatric disorders were significantly lower than in HC, whereas QA levels were not substantially different. | 6.3 | 23.8 |
| Chang et al.  (2017) | SI | 58 | 61 | MDD | Interview | Partly | Yes | Blood | - ESR | - ESR and hs-CRP levels in SI+ were significantly higher than in SI-. | 7.0 | 10.8 |
|  |  |  |  |  |  |  |  | Serum | - hs-CRP |  |  |  |
| Choi et al. (2021) | SI | 23 | 59 | MDD, Other | Self-report | Partly | Yes | Serum | - hs-CRP - IFN-γ - IL-10 - IL-6 - TNF-α | - Higher TNF-α levels were a significant indicator of SI in MDD patients. - No significant findings in SI+ patients with panic disorders. | 5.0 | 22.5 |
| De Beradis et al. (2014) | SI | 79 | 40 | Other | Self-report | Partly | Yes | Serum | - HDL | - No significant findings in SI+ patients with OCD. However, alexithymia patients with OCD showed significant lower HDL levels than HC. | 6.0 | 14.5 |
| Devici et al. (2007) | SA | 10 | 25 | Other | After SA | No | Yes | Serum | - BDNF | - Lower BDNF levels were observed in SA+ and MDD patients than in HC. | 5.0 | 22.5 |
| Dolsen et al. (2020) | SI | 244 | 1733 | Mixed | Self-report | No | Yes | Plasma | - hs-CRP - IL-6 - TNF-α | - Higher IL-6 levels were associated with SI+ in the past week. | 6.8 | 21.0 |
| Eidan et al. (2019) | SA | 22 | 34 | MDD | After SA | Partly | Yes | Plasma | - IL-6 - IFN-γ - HDL | - Higher IL-6 and IFN-γ levels were found in SA+ than in HC. - No significant difference in HDL was observed. | 7.5 | 14.5 |
| Eisen et al. (2016) | SA | 84 | 99 | Mixed | After SA | No | Yes | Serum | - BDNF | - No significant association between BDNF and SA. | 6.6 | 19.0 |
| Ekinci & Ekinci (2017) | SA | 37 | 76 | MDD | After SA | Partly | Yes | Serum | - HDL - hs-CRP | - NLR and hs-CRP levels in SA+ were significantly higher than in SA- or HC after adjusting the confounding factors. | 8.8 | 9.0 |
|  |  |  |  |  |  |  |  | Blood | - NLR |  |  |  |
| Fang et al. (2019) | SI | 26 | 148 | Other | Interview | Partly | Yes | Serum | - HDL | - No significant association between HDL and SI+ in schizophrenia patients. | 9.4 | 9.0 |
| Gabbay et al. (2009) | SA | 12 | 17 | MDD | After SA | Partly | Yes | Plasma | - IFN-γ - TNF-α - IL-1β - IL-6 - IFN-γ/IL-4 | - After adjusting confounding variables, IFN-γ levels in SA+ were significantly higher than in HC and TNF-α levels in SA+ were significantly lower than in SA-. | 4.8 | 17.0 |
| Ganança et al. (2021) | SI | 22 | 31 | MDD | Interview | Partly | No | Serum | - IL-1β - IL-6 - TNF-α | - Inflammatory and lipid markers revealed no significant differences in SI+ when compared with SI- or HC. - DHA% and IL-1β showed lower in patient with history of SA+ (within 5 years) when compared with SI+. | 5.0 | 15.0 |
|  |  |  |  |  |  |  |  | Plasma | - AA - EPA - DHA |  |  |  |
| Gibbs et al. (2016) | SA, SI | 129 | 110 | Mixed | After SA, Interview | No | No | Serum | - HDL - hs-CRP | - hs-CRP levels showed significantly higher in patient with SA+ when compared with SI+. - hs-CRP levels in SA+ were significantly higher than in SA- or HC. | 5.9 | 18.0 |
| Graça Cantarelli  et al. (2015) | SA | 50 | 36 | Affective | After SA | No | Yes | Serum | - BDNF - HDL | - No significant differences of BDNF and HDL between SA+ and SA-. | 5.9 | 19.3 |
| Grassi-Oliveira et al. (2012) | SI | 18 | 14 | MDD | Interview, Self-report | Yes | No | Plasma | - CCL2 - RANTES - CCL11 | - Lower CCL2 and RANTES levels were detected in SI+, compared with SA- and HC. | 4.8 | 12.0 |
| Grudet et al. (2014) | SA | 59 | 16 | Mixed | After SA | No | Yes | Serum | - Vitamin D | - Vitamin D levels in SA+ were significantly lower than in SA- or HC. | 4.1 | 21 |
| Gundogdu Meydaneri  et al. (2018) | SA | 27 | 26 | MDD | After SA | Partly | No | Blood | - WBC - NPs - Lymphs - Monocytes - NLR | - No significant differences of those biomarkers between groups. | 4.5 | 17.5 |
| Janelidze et al. (2015) | SA | 46 | 13 | Mixed | After SA | No | Yes | Plasma | - IL-8 | - No significant differences of IL-8 between SA+ and HC. | 6.3 | 17.3 |
| Khan et al. (2019) | SI | 28 | 58 | MDD | Interview | Yes | Yes | Serum | - BDNF | - BDNF levels in SI+ were significantly lower than in SI-. | 9.4 | 9.0 |
| Kim et al (2006) [a] | SA | 39 | 57 | MDD | After SA | Partly | Yes | Plasma | - NOx | - NOx levels in SA+ were significantly higher than in SA- or HC. | 8.4 | 16.0 |
| Kim et al. (2007) [b] | SA | 32 | 31 | MDD | After SA | Partly | No | Plasma | - BDNF | - BDNF levels in SA+ were significantly lower than in SA- or HC. | 4.7 | 17.5 |
| Kim et al. (2014) [c] | SI | 93 | 639 | Not Applicable | Interview | No | Yes | Serum | - HDL | - No significant differences of HDL between groups. | 5.3 | 21.5 |
| Kurosawa et al. (2018) | SA | 33 | 146 | Mixed | After SA | No | No | Plasma | - AA - EPA - DHA | - EPA levels were negatively associated with SA+, whilst DHA levels were positively associated. | 4.4 | 19.5 |
| Lee at al. (2006) [a] | SA | 53 | 67 | Mixed | After SA | Partly | Yes | Plasma | - NOx | - NOx levels in SA+ were significantly higher than in SA- or HC. | 8.1 | 15.5 |
| Lee et al. (2007) [b] | SA | 28 | 72 | MDD | After SA | Partly | Yes | Plasma | - BDNF | - BDNF levels in SA+ were significantly lower than in SA-. | 9.7 | 10.5 |
| Liu et al. (2020) | SI | 24 | 61 | MDD | Self-report | Yes | No | Serum | - IL-1β - IL-6 - IFN-γ - TNF-α - Chemokines (i.e., CCL8, IL-8, etc.) | - CCL8 levels in SI+ were significantly higher than in SI- or HC. - No further significant finding was observed between SI+ and SI- or HC. | 7.0 | 9.8 |
| Loas et al. (2016) | SA | 41 | 81 | Mixed | After SA | No | Yes | Serum | - hs-CRP | - hs-CRP levels in SA+ were significantly higher than in SA-. | 3.0 | 19.0 |
| Ma et al. (2020) | SA | 235 | 1372 | MDD | Interview | Partly | Yes | Plasma | - HDL | - HDL levels in SA+ were significantly lower than in SA-. | 5.9 | 17.5 |
| Maes et al. (1989) | SI | 17 | 17 | MDD | Interview | Partly | Yes | Blood | - TRP - TRP/CAA | - No significant differences between SI+ and SI- in TRP or TRP/CAA. | 4.7 | 13.5 |
| Mensi et al. (2016) | SA | 15 | 71 | Other | After SA | Partly | Yes | Serum | - HDL | - HDL had no significant association with SA in schizophrenia patients. | 4.1 | 22.5 |
| Mohamed et al. (2020) | SI | 24 | 74 | MDD | Self-report | Partly | No | Serum | - hs-CRP | - hs-CRP levels in SI+ were significantly higher than in SI-. | 4.1 | 18.5 |
| Nässberger & Träskman-Bendz(1993) | SA | 30 | 25 | Mixed | After SA | Partly | No | Plasma | - sIL-2R | - sIL-2R levels in SA+ were significantly higher than in HC. | 4.5 | 21.5 |
| O’Donovan  et al. (2013) | SI | 29 | 48 | MDD | Interview | Partly | Yes | Plasma | - hs-CRP - IL-6 - IL-10 - TNF-α | - IL-6 and hs-CRP levels in SI+ were significantly higher than in HC. | 6.0 | 21.0 |
| Oh et al. (2020) | SA | 405 | 3584 | Affective | Interview | No | No | Blood | - hs-CRP - ESR - WBC - Vitamin D | - hs-CRP and ESR levels in SA+ were significantly higher than in SA-. | 1.6 | 25 |
| Oshnokhah  et al. (2020) | SA | 50 | 40 | Excluded psychiatric disorders group | After SA | Yes | Yes | Serum | - MDA - NOx - TAC | - MDA and TAC levels were substantially lower in SA+ than in HC, whereas NOx levels were significantly higher. | 6.9 | 14.0 |
| Özer et al. (2004) | SI | 10 | 17 | Other | Interview | Partly | Yes | Serum | - HDL | - No significant association between HDL and SI+ in panic patients. | 5.9 | 14.0 |
| Park et al. (2014) | SI | 18 | 56 | MDD | Interview, Self-report | No | Yes | Serum | - HDL | - No significant differences of HDL between groups. | 5.0 | 18.5 |
| Pasyk et al. (2020) | SA | 117 | 176 | Mixed | After SA | No | Yes | Serum | - BDNF | - BDNF levels were shown to be significantly associated with self-reported impulsivity scores, but not with SA. | 6.6 | 19.0 |
| Peng et al. (2018) | SA | 69 | 202 | MDD | After SA | Partly | Yes | Serum | - hs-CRP - HDL - Albumin | - No significant differences among those biomarkers between SA+ and SA-. | 8.8 | 14.0 |
| Pinheiro et al. (2012) | SI | 14 | 176 | Affective | Interview | No | Yes | Serum | - BDNF | - BDNF levels were significant lower in SI+ than in SI-. | 7.2 | 21.5 |
| Priya et al. (2016) | SA | 42 | 42 | Not Applicable | After SA | No | No | Serum | - BDNF - hs-CRP - IL-6 | - BDNF levels were substantially lower in SA+ than in HC, whereas hs-CRP and IL-6 levels were significantly higher. - After adjusting for confounder factors, linear regression indicated hs-CRP as a predictor of suicide risk. | 5.6 | 18.0 |
| Rasheed et al. (2019) | SA | 22 | 34 | MDD | After SA | Partly | Yes | Plasma | - HDL - IL-1β - TNF-α | - TNF- levels were significantly higher in SA+ than in SA- or HC, while there were no significant differences in IL-6 or HDL levels between the groups. | 6.0 | 12.0 |
| Roy & Roy (2006) | SI | 61 | 397 | Affective | Self-report | No | No | Serum | - HDL | - No significant association between HDL and SI+ in depressive patients with Type I Diabetes. | 2.8 | 24.5 |
| Ruljancic et al. (2013) | SA | 79 | 89 | MDD | After SA | No | No | Serum | - Albumin | - Albumin levels were significantly lower in SA+ than in SA- or HC. | 5.3 | 18.4 |
| Segoviano-Mendoza et al. (2018) | SA | 59 | 204 | MDD | After SA | Partly | Yes | Serum | - HDL | - No significant differences of HDL between groups. | 7.5 | 16.5 |
| Si et al. (2020) | SI | 241 | 441 | Not Applicable | Self-report | No | Yes | Serum | - HDL | - No significant differences of HDL between SI+ and SI-. - Females with SI had higher HDL concentrations than males with SI, but females and males without SI revealed no significant differences. | 4.0 | 20.5 |
| Sowa-Kucma et al. (2018) | SI | 93 | 95 | Affective | Interview | Partly | Yes | Serum | - zCMI+TBARS - sIL-1RA - sIL-2R - sIL-6R - IL-1α - TBARS - sTNFR60 - sTNFR80 | - Increased TBARS was associated with SI. - In SI, there were no significant differences in other biomarkers across groups. | 9.3 | 9.0 |
| Su et al. (2019) | SI | 143 | 219 | Affective | Self-report | No | No | Serum | - HDL | - HDL levels were significant higher in SI+ than in SI-. | 5.3 | 20.5 |
| Velasco et al. (2020) | SA | 126 | 136 | MDD | Interview | Yes | Yes | Blood | - Leukocytes - Lymphs - Monocytes - NPs - NLR - MLR | - Lymphs and NLR were significant differences between SA+ and SA-, whilst there were no significant differences in other biomarkers across groups. | 5.5 | 16.0 |
| Verma et al. (1999) | SA | 40 | 40 | Mixed | After SA | No | Yes | Serum | - HDL | - HDL levels revealed no significant differences between SA+ and SA-. | 5.0 | 12.5 |
| Yagci & Avci (2021) | SA | 40 | 41 | Mixed | After SA | Partly | Yes | Blood | - CRP - Leukocytes - NLR | - CRP, leukocytes, and NLR concentrations were significant higher in SA+ than in HC. | 3.8 | 12.5 |

**Abbreviations:**

AA: Arachidonic acid (omega-6 fatty acid)

BDNF: Brain-derived neurotrophic factor

CCL: The C–C motif chemokine ligand

CRP: C-reactive protein

DHA: Docosahexaenoic acid (omega-3 fatty acid)

EPA: Eicosapentaenoic acid (omega-3 fatty acid)

HDL-c: High-density lipoprotein cholesterol

hsCRP: The high-sensitivity C-reactive protein

IL: Interleukin

IFN: Interferon

KYN: Kynurenine

Lymphs: Lymphocytes

MDA: Malondialdehyde

MLR: Monocyte to lymphocyte ratio

NLR: Neutrophil-to-lymphocyte ratio

NOx: Nitric oxide metabolites

NPs: Neutrophils

PA: Picolinic acid

QA: Quinolinic acid

RANTES: Regulated on Activation, Normal T Cell Expressed and Secreted

SA: Suicide attempts

SI: Suicide ideation

sIL2R: Soluble Interleukin 2 Receptor

sIL6R: Soluble Interleukin 6 Receptor

sTNF-αR1: Soluble tumor necrosis factor alpha receptor 1

TAC: Total antioxidant capacity

TBARS: Thiobarbituric acid reactive substances

TNF: Tumor necrosis factor

TNFR: Tumor necrosis factor receptors

TRYCATs: The tryptophan catabolites

TRP: Tryptophans

**ESF 1 Table 5.** Excluded studies.

| **Authors, year** | **Reason why excluded** | **Key findings** |
| --- | --- | --- |
| Dickerson et al., 2017^a^ | Graph format. | There was a significantly higher level of CRP in the patients with history of suicide attempts more than 1 month (coefficient=0.87, 95% CI 0.25, 1.50, p=0.006) compared with the control group but not in the other two psychiatric groups (p > 0.05), including patients with history of suicide attempts in 1 month and without history of suicide attempts, when adjusting for age, gender, race, smoking status, and BMI |
| Jha et al., 2020^b^ | Combined case group with suicide attempts and suicidal ideation. | After adjustment, only IL-4 revealed significantly differences between healthy control (n=39), those at risk of MDD group (n=33), and those at risk of MDD with recent suicide attempts or suicidal ideation group (n=37), whereas the other cytokines and chemokines did not. Plasma IL-4 levels of recent suicide group had lower than healthy controls and at risk of MDD group. |
| Lauterbach et al., 2006^c^ | Correlation coefficient. | There is a modest correlation between suicidal ideation and low plasma trytophan/amino acid ratio (*r*=0.39, p=0.042). |
| Melhem et al., 2017^d^ | Logarithmic trasformation. The authors did dot provide mean (SD) values upon request. | Patients with suicide attempts showed significantly higher CRP [95% CI (0.15, 1.84), p = 0.02] compared to patients with suicidal ideation and healthy controls. |
| Roggenbach et al., 2007^e^ | Combined case group with suicide attempts and suicidal ideation. | There is no significant different between depression patients with suicide group and healthy control in plasma tryptophan. |

a. Dickerson F, Adamos M, Katsafanas E, Khushalani S, Origoni A, Savage C et al. The association between immune markers and recent suicide attempts in patients with serious mental illness: A pilot study. Psychiatry Res 2017; **255**: 8-12.
b. Jha MK, Cai L, Minhajuddin A, Fatt CC, Furman JL, Gadad BS et al. Dysfunctional adaptive immune response in adolescents and young adults with suicide behavior. Psychoneuroendocrinology 2020; **111**: 104487.
c. Lauterbach E, Brunner J, Hawellek B, Lewitzka U, Ising M, Bondy B et al. Platelet 5-HT2A receptor binding and tryptophan availability in depression are not associated with recent history of suicide attempts but with personality traits characteristic for suicidal behavior. Journal of Affective Disorders **2006**; 91(1): 57-62.
d. Melhem NM, Munroe S, Marsland A, Gray K, Brent D, Porta G et al. Blunted HPA axis activity prior to suicide attempt and increased inflammation in attempters. Psychoneuroendocrinology 2017; **77**: 284-294.
e. Roggenbach J, Müller-Oerlinghausen B, Franke L, Uebelhack R, Blank S, Ahrens B. Peripheral serotonergic markers in acutely suicidal patients. 1. Comparison of serotonergic platelet measures between suicidal individuals, nonsuicidal patients with major depression and healthy subjects. Journal of Neural Transmission 2007; **114**(4): 479-487.
