## Supplementary material for "Inflammation and nitro-oxidative stress in current suicidal attempts and current suicidal ideation: a systematic review and meta-analysis": ESF 2

**Electronic Supplementary Information File (ESF) 2**

Activated immune, oxidative and nitrosative stress pathways are strongly associated with suicidal attempts and less with suicidal ideation: a meta-analysis and meta-regression.

**Running title:** immune activation in suicidal behaviors

Asara Vasupanrajit, M.Sc.^a^; Ketsupar Jirakran, M.Sc.^a,b^; Chavit Tunvirachaisakul, M.D., Ph.D.^a,^ ^c^; Michael Maes, M.D., Ph.D.^a, c, d, e^

^a^ Department of Psychiatry, Faculty of Medicine, Chulalongkorn University, Bangkok, Thailand.

^b^ Maximizing Thai Children’s Developmental Potential Research Unit, Department of Pediatrics, Faculty of Medicine, Chulalongkorn University, Bangkok, Thailand.

^c^ Cognitive Impairment and Dementia Research Unit, Department of Psychiatry, Faculty of Medicine, Chulalongkorn University, Bangkok, Thailand.

^d^ IMPACT Strategies Research Center, Deakin University, Geelong, Australia.

^e^ Department of Psychiatry, Medical University of Plovdiv, Plovdiv, Bulgaria.

**ESF 2 Table 1.** Results of other subgroup analyses performed on IO&NS profile.

| **Comparison** | **n studies** | **subgroups** | **SMD** | **95% CI** | **z** | **p** | **Q** | **df** | **p** | **I^2^  (%)** | **τ^2^** | **Τ** |
| --- | --- | --- | --- | --- | --- | --- | --- | --- | --- | --- | --- | --- |
| **Subgroups by psychiatric groups** | | | | | | | | | | | | |
| SB *versus* Controls (χ^2^ =8.563; df=3; p=0.036) | 29 | MDD | 0.370 | 0.195; 0.546 | 4.135 | <0.001 | 141.577 | 28 | <0.001 | 80.223 | 0.168 | 0.410 |
|  | 15 | MIX | 0.372 | 0.201; 0.543 | 4.267 | <0.001 | 57.569 | 14 | <0.001 | 75.681 | 0.075 | 0.274 |
|  | 9 | Affective | 0.049 | -0.156; 0.254 | 0.470 | 0.639 | 30.696 | 8 | <0.001 | 73.938 | 0.063 | 0.251 |
|  | 6 | OTHER | 0.140 | -0.067; 0.346 | 1.326 | 0.185 | 3.577 | 5 | 0.612 | 0.000 | 0.000 | 0.000 |
| **Subgroups by participants groups** | | | | | | | | | | | | |
| SB *versus* Controls  (χ^2^ = 8.147; df=1; p=0.004) | 33 | In-patient | 0.442 | 0.264; 0.620 | 4.862 | <0.001 | 203.090 | 32 | <0.001 | 84.243 | 0.215 | 0.464 |
|  | 18 | Out-patient | 0.128 | 0.007; 0.249 | 2.080 | 0.038 | 37.804 | 17 | 0.003 | 55.031 | 0.027 | 0.165 |
| SA *versus* Controls  (χ^2^ = 5.770; df=1; p=0.016) | 26 | In-patient | 0.505 | 0.312; 0.698 | 5.126 | <0.001 | 137.711 | 25 | <0.001 | 81.846 | 0.193 | 0.439 |
|  | 5 | Out-patient | 0.151 | -0.064; 0.366 | 1.375 | 0.169 | 12.688 | 4 | 0.013 | 68.474 | 0.031 | 0.177 |
| **Subgroups by medium** | | | | | | | | | | | | |
| SB *versus* Controls  (χ^2^ =4.677; df=2; p=0.096) | 7 | Blood | 0.229 | 0.066; 0.391 | 2.759 | 0.006 | 10.229 | 6 | 0.115 | 41.341 | 0.018 | 0.132 |
|  | 19 | Plasma | 0.509 | 0.275; 0.742 | 4.270 | <0.001 | 123.125 | 18 | <0.001 | 85.381 | 0.199 | 0.446 |
|  | 39 | Serum | 0.232 | 0.117; 0.347 | 3.958 | <0.001 | 147.864 | 38 | <0.001 | 74.301 | 0.089 | 0.298 |
| **Subgroups by the assessment of suicide behaviors** | | | | | | | | | | | | |
| SB *versus* Controls  (χ^2^ = 8.479; df=2; p=0.014) | 32 | Suicidal act | 0.456 | 0.291; 0.622 | 5.409 | <0.001 | 155.948 | 31 | <0.001 | 80.122 | 0.168 | 0.410 |
|  | 18 | Interview | 0.173 | 0.070; 0.275 | 3.304 | 0.001 | 28.494 | 17 | 0.039 | 40.338 | 0.016 | 0.126 |
|  | 11 | Self-report | 0.184 | -0.031; 0.400 | 1.676 | 0.094 | 58.554 | 10 | <0.001 | 82.922 | 0.096 | 0.310 |
| SA *versus* Controls  (χ^2^ =7.102; df=1; p=0.008) | 32 | Suicidal act | 0.456 | 0.291; 0.622 | 5.409 | <0.001 | 155.948 | 31 | <0.001 | 80.122 | 0.168 | 0.410 |
|  | 5 | Interview | 0.120 | -0.064; 0.304 | 1.277 | 0.201 | 13.129 | 4 | 0.011 | 69.533 | 0.026 | 0.161 |
| SI *versus* Controls  (χ^2^ = 0.015; df=1; p=0.902) | 13 | Interview | 0.200 | 0.069; 0.331 | 3.003 | 0.003 | 15.362 | 12 | 0.222 | 21.886 | 0.012 | 0.110 |
|  | 11 | Self-report | 0.184 | -0.031; 0.400 | 1.676 | 0.094 | 58.554 | 10 | <0.001 | 82.922 | 0.096 | 0.310 |

SMD: standardized mean difference, 95% CI: 95% confidence intervals

IO&NS: immune-inflammatory and oxidative and nitrosative stress

SB: suicide behaviors

SA: suicide attempts

SI: suicidal ideation

We used the prespecified subgroups as the units of analysis. Thus, patients with suicide behaviors were first compared with controls. Second, we compared a) SA+ *versus* controls); and b) SI+ *versus* controls.

**ESF 2 Table 2.** Results on publication bias.

| **Outcome feature sets** | **Fail safe n** | **Z Kendall’s τ** | **p-value  (1-tailed)** | **Egger’s t test (df)** | **p-value  (1-tailed)** | **Missing studies (side)** | **Adjusted SMD (95%CI)** |
| --- | --- | --- | --- | --- | --- | --- | --- |
| IO&NS | 1829 | 1.44 | 0.075 | 2.50 (57) | 0.008 | 6 (R) | 0.371 (0.263; 0.479) |
| IRS | 1304 | 0.73 | 0.233 | 2.67 (31) | 0.006 | 5 (R) | 0.551 (0.403; 0.710) |
| Inflammation | 828 | 1.06 | 0.145 | 2.07 (24) | 0.025 | 3 (R) | 0.556 (0.376; 0.737) |
| CRP | 529 | 2.57 | 0.005 | 3.24 (13) | 0.003 | 2 (R) | 0.692 (0.428; 0.956) |
| Neurotoxicity | 776 | 0.13 | 0.448 | 1.93 (27) | 0.032 | 4 (R) | 0.490 (0.335; 0.645) |
| ANTIOXPRO | 319 | 0.72 | 0.235 | 1.03 (38) | 0.154 | 4 (R) | 0.249 (0.124; 0.374) |
| BDNF | 110 | 0.89 | 0.186 | 1.99 (8) | 0.041 | 0 | - |

SMD: standardized mean difference, 95% CI: 95% confidence intervals

IO&NS: immune-inflammatory and oxidative and nitrosative stress

IRS: immune-inflammatory response system

ANTIOXPRO: protection via antioxidants and neurotrophic products.

CRP: C-reactive protein

BDNF: Brain-derived neurotrophic factor

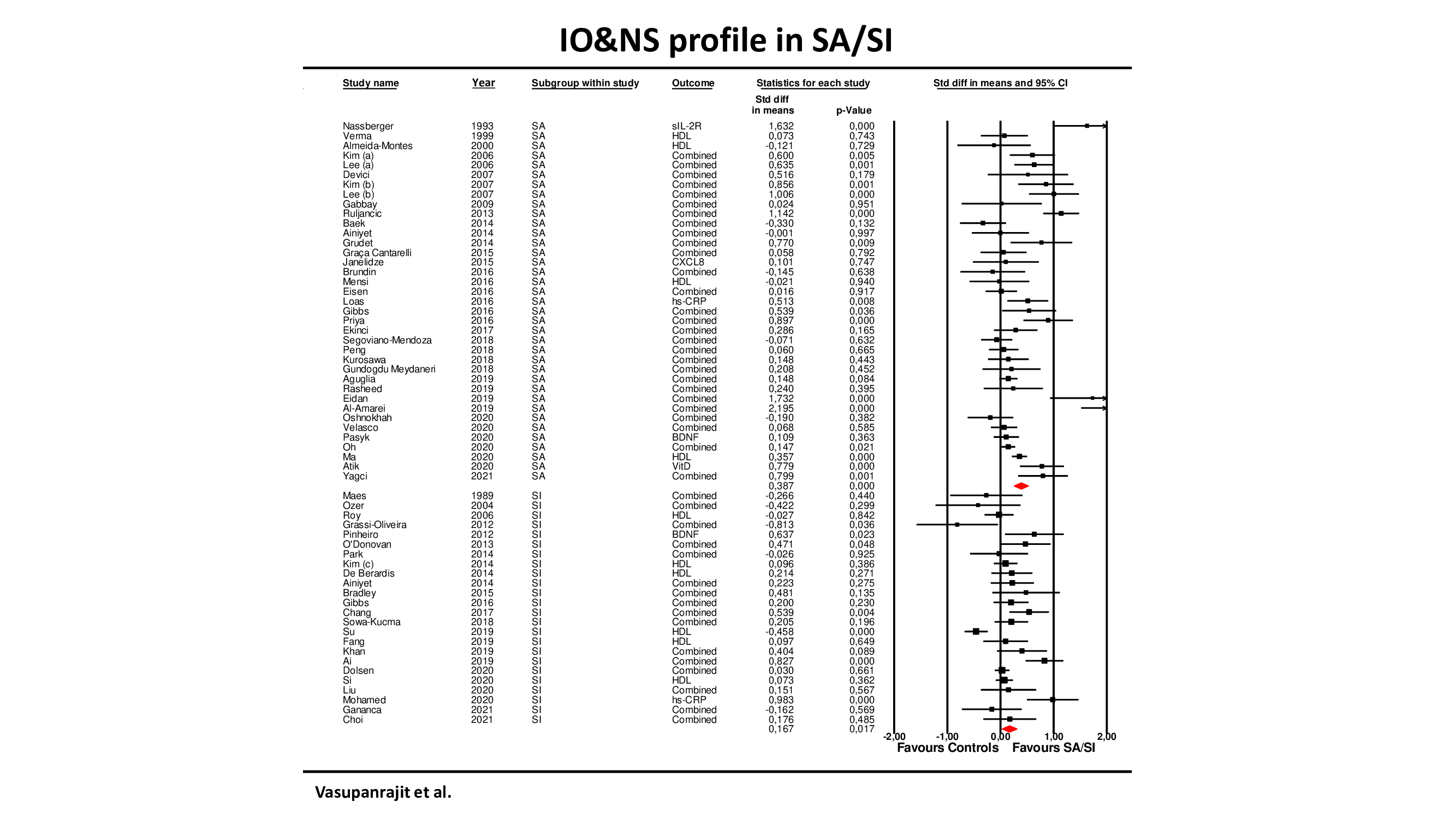

**ESF 2 Figure 1.** Forest plot with results of subgroup analysis performed on 61 suicide attempts (SA) or suicidal ideation (SI) studies reporting immune-inflammatory and oxidative & nitrosative stress (IO&NS) biomarkers.

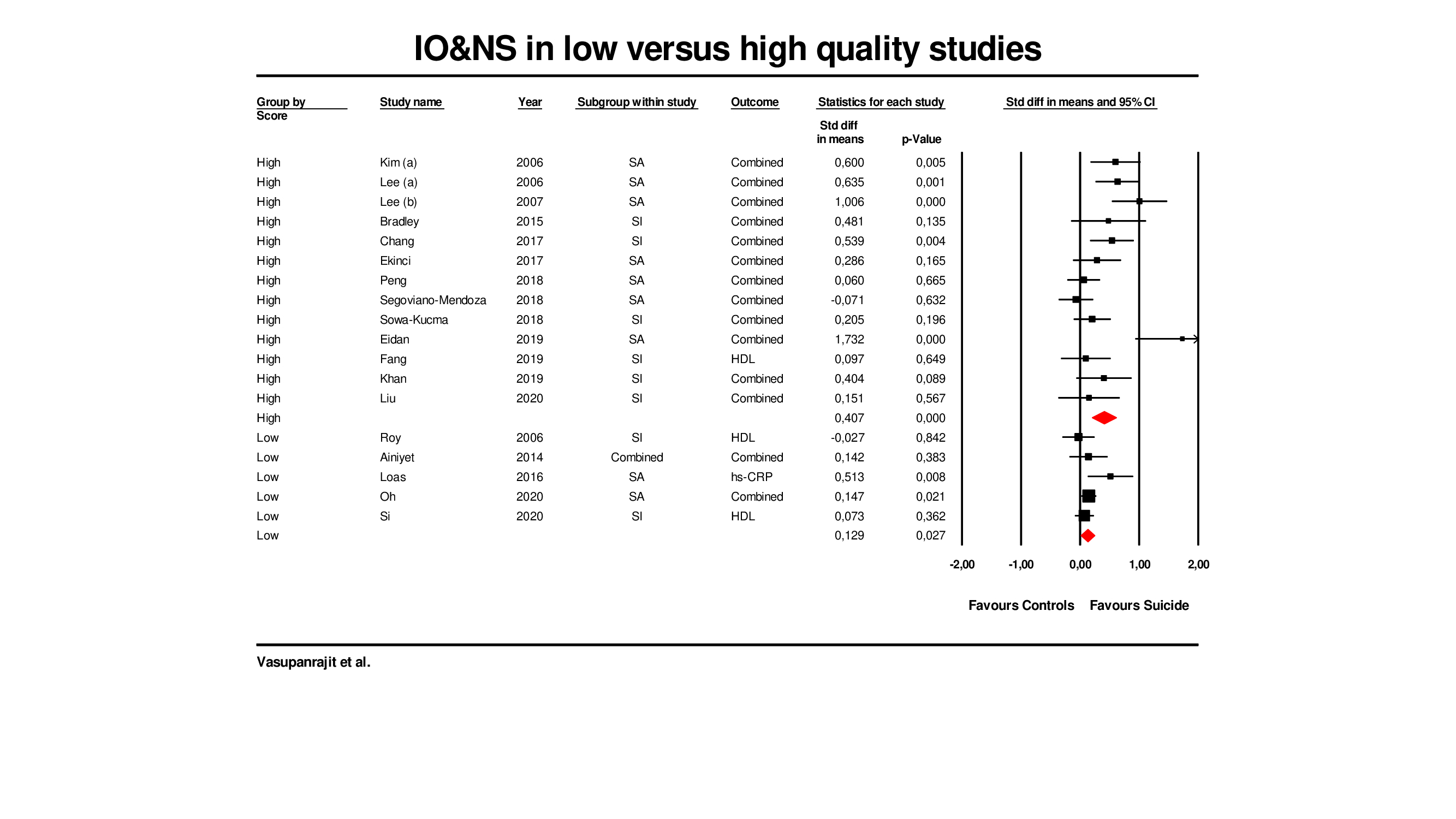

**ESF 2 Figure 2.** Forest plot with results of meta-analysis performed on 13 high- and 5 low-quality studies reporting immune-inflammatory and oxidative & nitrosative stress (IO&NS) biomarkers when comparing suicide behaviors to controls.
